# Genome-wide association studies of social participation and occupational engagement in the UK Biobank

**DOI:** 10.1101/2025.05.15.25327674

**Authors:** Evie Doherty, Aodán Laighneach, Mia Casburn, Fergus Quilligan, Gary Donohoe, Dara M. Cannon, Derek W. Morris

**Affiliations:** Centre for Neuroimaging, Cognition and Genomics (NICOG), University of Galway, Ireland; School of Biological and Chemical Sciences, University of Galway, Ireland; Clinical Neuroimaging Laboratory, School of Medicine, University of Galway, Ireland; School of Psychology, University of Galway, Ireland

**Keywords:** Psychosis, Psychosocial Disability, FastGWA, Schizophrenia

## Abstract

Psychosis is a clinically heterogenous disorder associated with significant difficulties with social and occupational function (psychosocial disability; PD). While environmental and cognitive factors are identified predictors of PD, the genetic contribution remains unclear. Here, we investigated the hypothesis that objective social participation (SP) and occupational engagement are genetically influenced.

We performed mixed-linear-model genome-wide association studies of these phenotypes in the UK Biobank (*N*∼404,500) and a series of post-hoc analyses including Mendelian randomization (MR) to interpret findings. SP was defined as the frequency of social visits and leisure activities based on response to questionnaires. Occupational engagement was represented by two variables; occupational function (OF) and the established Not in Education, Employment, and Training (NEET) measure, both derived from employment status responses. We identified 17 independent loci for SP, with a SNP-based heritability of 4.1%. A list of contributory genes included *CSE1L*, *TNRC6B*, *STAU1, CDH7*, *GBE1*, *ZNF536, DDX27*, and the known schizophrenia risk gene, *TCF4*. The regulation of synaptic signalling was implicated in the biology of SP by gene-set analysis. SNP-based heritabilities for OF and NEET were 1.8% and 1.2% respectively and *DRD2* was associated with both phenotypes. Reduced SP and occupational engagement demonstrated genetic correlations with an increased risk for neuropsychiatric disorders, socioeconomic deprivation, lower cognitive ability, loneliness, neuroticism and chronic pain. MR indicated that attention-deficit hyperactivity disorder and schizophrenia were causal for reduced occupational engagement. PD has a genetic component with shared genetic links and relationships with neuropsychiatric disorders and related traits.

## Introduction

Psychosocial disability (PD) refers to the social and occupational challenges associated with mental ill-health.^1^ These challenges are considered distinct from psychiatric disability, which encapsulates clinical impairment. Although a common impairment in neuropsychiatric disorders,^2–5^ there is little consensus on how to define social and occupational function in the literature.^6^ Here, we define social participation (SP) as the degree to which an individual actively participates in social activities and/or interactions, and occupational engagement as the extent to which an individual actively engages occupationally/vocationally. We aimed to objectively capture how well a person actively functions in society rather than how a person subjectively views their function.

PD is widely reported in psychosis and related disorders including schizophrenia (SCZ) and bipolar disorder (BD).^5,7^ Although psychosis is generally characterised by positive, negative, and cognitive symptoms, deficits in social and occupational function are also key features. Psychosis is a leading cause of burden in terms of years lived with disability worldwide^8^ and has a lifetime prevalence of ∼3%.^9^ Often present prior to symptom onset, psychosocial difficulties have been linked to both development^10–12^ and trajectory^13,14^ of psychosis, making social and occupational function potentially valuable early intervention targets.

Community detachment, education and hobby avoidance, and impairment in work-related activities are common psychosocial challenges in psychosis.^15^ Even during periods of clinical remission, up to 50% of individuals with psychosis fail to regain full psychosocial functioning.^16,17^ This reflects the fact that current pharmacological interventions have scant effects on social and occupational deficits. Factors such as cognitive performance,^18,19^ adverse life events,^20^ duration of untreated psychosis,^21^ and sociodemographic factors significantly contribute to PD in psychosis. However, functional deficits remain understudied and the biological underpinnings of SP and occupational engagement remain unclear. Understanding the genetic determinants of SP and occupational engagement is a fundamental step in the prediction of functional outcomes and in overcoming the persistent problem of poor functional recovery in psychosis.

Several genome-wide association studies (GWAS) have explored the genetics of social phenotypes including social interaction,^22^ loneliness,^22,23^ sociability,^24^ and social-isolation.^25^ In the largest GWAS to date, 17 independent genomic regions were linked to social-isolation,^25^ implicating *ARFGEF2*, *ZNF536*, *CSE1L*, and *DIAPH3*, genes associated with social deficit disorders including SCZ^26^ and attention deficit hyperactivity disorder (ADHD).^27,28^ Despite other studies also linking social phenotypes to genes implicated in these disorders,^22,24^ there is little homogeneity in results. In the UKB, GWASs for specific domains of social functioning have been performed. For frequency of friend/family visits, lead SNPs near genes associated with social functioning phenotypes were identified,^24,25^ as well as several novel genes. GWASs were also carried out for leisure activity type (e.g. Sports Club/Gym), however, results had little concordance with previous research. Research to date has been largely unsuccessful in identifying robust genetic variants associated with social functioning, likely due to the varying interpretations of social function.

GWASs of occupational function (OF) are limited. Research focuses on socioeconomic status (SES) to investigate the genetic determinants of OF and proxy phenotypes such as educational attainment^29,30^ and income^31,32^ neglecting occupational status. Twin studies show that heritability of occupational status (35- 45%)^33^ is comparable to that of educational attainment (∼40%) and income (∼40%).^33,34^ To date, just two GWASs of occupational status have been conducted. The first carried out individual GWASs for each current employment type in the UKB^23^ (e.g. employed/unemployed/retired), identifying eight lead-SNPs. The most recent study identified 106 variants.^35^ Further analyses revealed a strong genetic correlation between occupational status, educational attainment, and income despite evidence that occupational status may be somewhat empirically distinct.

In the present study, we conducted three GWASs of SP and occupational engagement phenotypes in >404,000 participants from the UKB using a mixed linear methods approach.^36^ We performed SNP-based heritability and explored genetic correlations with ∼1,500 traits and psychiatric disorders. We integrated multiple functional genomics datasets, performed gene prioritisation and transcriptome-wide association analysis to identify susceptibility genes. Finally, we implemented Mendelian randomization, investigating the casual relationships between our phenotypes and associated traits and disorders.

## Methods

### Study Design and Population

This was a large-scale, cross-sectional analysis of UKB participants. The UKB is a biomedical database containing extensive genetic and phenotypic data for 502,292 individuals aged 37-73 years from across the United Kingdom. Participants were recruited from 23 assessment centers during the period of 2006-2010, capturing rural and urban demographics with heterogenous ethnic and socioeconomic backgrounds. The average age at recruitment was 56.5 years with female sex more common (54%). This research was conducted under the UKB application number 98153 and follows UKB Ethics and Governance Framework.

### Phenotypes

SP was constructed by creating a composite score from two UKB database questions that capture objective social activity and interaction; (1) a question on the number of leisure/social activities attended weekly (Data-Field 6160), and (2) a question on frequency of friend/family visits (Data-Field 1031). Possible scoring ranged from 0 to 10. Occupational engagement was represented by two individual variables: OF and Not in Education, Employment, and Training (NEET) status, a pre-established variable from youth mental health research.^37^ Both variables were derived from the UKB database question on current employment status (Data-Field 6142). Scoring ranged from -2 to 4 for OF, while NEET status was binary (0=non-NEET, 1=NEET; see Supplementary Methods).

### Phenotypic Data on Disorders of Interest

Individuals with SCZ, BD, major depressive disorder (MDD), and ADHD were grouped by ICD-10 diagnosis codes. The unaffected group was created by removing all individuals with any of the above ICD-10 diagnoses. R software (v4.3.1)^38^ was used to calculate mean scores and distributions and perform general linear models to compare neuropsychiatric and the unaffected groups (correcting for age, sex and assessment center; see Supplementary Methods).

### Genetic Data

The genotype dataset made available by the UKB in the December 2023 release was used. Genetic data was genotyped using the Applied Biosystems UK Biobank Axiom Array^39^ or the Applied Biosystems UK BiLEVE Axiom Array^40^ (see Supplementary Methods). Genotypes were imputed as described previously.^41^ Quality control (QC) was carried out on imputed data, filtering variants with a minor allele frequency (MAF) >0.0001 and an imputation quality information (INFO) score >0.9, reducing the number of usable SNPs to 15,857,586. Individuals with discordant sex, sex aneuploidies, or of non-white British ancestry were removed with further QC.

### Statistical Analyses

GWASs of SP, OF, and NEET status were performed with fastGWA^36^ using genome-wide complex trait analysis (GCTA) software (v1.94.1). FastGWA maximizes power through a genetic relationship matrix that allows the inclusion of related individuals within analyses (see Supplementary Methods). The FastGWA default QC process removed variants with >10% missingness. GWASs were performed adjusting for age (Data Field 21003), sex (Data Field 31), array (Data Field 22000), Townsend Deprivation Index (TDI) scores (Data Field 22189), assessment center (Data Field 54) and the first 20 principal components.

### SNP-based Heritability

LD score regression (LDSC) (v1.0.1) was applied to estimate the SNP-based heritability from GWAS summary statistics.^42^ Pre-computed LD scores from the 1000 Genomes Project European samples were used to carry out the analysis (https://github.com/bulik/ldsc).

### Statistical Fine-mapping

To narrow the credible window of the risk loci and identify potentially causal variants associated with SP and OF, sum of single effects (SuSiE)^43^ was used with the 1000 Genomes Project European reference panel (Phase 3).^44^ Credible sets were identified using the susieR package (v0.12.35)^45^ in R (v4.4.0)^38^ (see Supplementary Methods). The posterior inclusion probability (PIP) score (>0.5) was used to determine the likely causal variant at a given region.

### Functional Annotation

FUMA online pipeline (v1.5.2)^46^ was used to map and annotate fastGWA outputs. Annotation was performed for all nominally significant SNPs and any variant in LD (r^2^≥0.6). ANNOVAR was used to identify genic positions of SNPs, while Combined Annotation Dependent Depletion (CADD) scores were generated to determine variant pathogenicity (CADD score >12.37 = potentially pathogenic).

### Gene-based Analysis

Gene-based analysis was carried out using Multi-marker Analysis of GenoMic Annotation (MAGMA) (v1.6.) within the Functional Mapping and Annotation (FUMA) online tool (v1.5.2).^46^ Gene-set analyses were implemented using 10,678 gene sets from MsigDB (v6.2) (curated gene-sets: 4,761; GO terms: 5,917). A Bonferroni correction threshold of *P*<2.63e-6 was applied (0.05/19036 protein coding genes).

### Gene-mapping

Genome-wide significant loci were mapped to genes using the FUMA online tool (v1.5.2)^46^ using positional, eQTL, and 3D chromatin interaction mapping strategies (see Supplementary Methods).

### Gene Prioritization

To further annotate potentially functional genes associated with the phenotype-specific variants, Polygenic Priority Score (PoPS) was performed using 57,543 gene-based features from 77 gene expression datasets.^47^ PoPS is a similarity-based gene prioritization method used to identify causal genes at GWAS loci. Gene-level association statistics and gene–gene correlations were computed using MAGMA (v1.0.1)^48^ and LD estimates from the European panel of the 1000 Genomes Project (Phase 3). PoPS were computed for remaining genes (*N*=18,224) which were then prioritized by selecting the highest scoring gene within 500-kb, either direction, of the GWS variants at each locus in R (v4.3.1).^38^

### Transcriptome-wide association study

A transcriptome-wide association study (TWAS) analysis was conducted using precomputed gene expression weights from 1,321 PsychENCODE postmortem brain samples^49^ (http://resource.psychencode.org/). FUSION software^50^ was used to examine whether SNPs influencing gene expression (12,041 genes) were associated with SP and OF (*P*bon <4.19e-06), (see Supplementary Methods). TWAS fine-mapping was performed using FOCUS (v0.09)^51^ in regions including a TWAS significant gene (see Supplementary Methods).

### Genetic Correlation Analyses

Bivariate genetic correlations were calculated between our UKB phenotypes and summary statistics for SCZ^52^, BD^53^, MDD^54^, ADHD^55^, and Autism Spectrum Disorder (ASD)^56^, from the Psychiatric Genomics Consortium using LDSC (v1.0.1). A batch genetic correlation of 1,583 phenotypes was also performed using the Complex-Traits Genetics Virtual Lab (CTG-VL)^57^ (*P-*value threshold <1.39e-5).

### Mendelian Randomization

Mendelian randomization (MR) was conducted using the TwoSampleMR R package (v0.6.8).^58^ Instrumental variables were genome-wide significant (GWS) SNPs reported in GWAS of European ancestry samples only and minus UKB samples (SP, SCZ^52^, BD^53^, MDD^59^, ADHD^55^, ASD^56^, and educational attainment (EA)^29^ using the statistical significance threshold of *P*<5x10e-8. OF, NEET and ASD were not considered as exposures because of too few GWS SNPs. EA was not considered as an outcome because full GWAS results were not available for a European ancestry sample excluding UKB. The MR-Egger method was used for analyses to account for potential horizontal pleiotropy (see Supplementary Methods). The *P*_bon_ threshold was determined by the number of tests, 0.05/20 tests = 0.0025.

## Results

### UKB SP and OF and NEET demographics

Of the 487,409 UK Biobank participants with genotype data, 404,403 (54.2% female, mean age 56.9 years, sd=7.99) were available for GWAS analysis of SP after filtering and QC was performed. The SP scores ranged from 0 (low SP) to 7 (high SP), with a population mean of 4.28. For OF and NEET status variables, 404,569 (54.2% female, mean age 56.9 years, sd=8.00) and 404,469 individuals (54.2% female, mean age 56.9 years, sd = 8.00), respectively, were included in the final GWAS analyses. Scoring for OF ranged from -2 to 4, with 59.1% attaining a score of 1 (mean score 0.61). NEET status was scored simply as 1 (non-NEET) or 2 (NEET), of which 42.9% reported being of NEET status.

### Diagnostic comparison of mean phenotypic scores

Mean phenotypic measure scores of participants from psychiatric disorder groups (SCZ n= 1,659, BPD n=1,544, MDD n=27,276) were each compared to those of the unaffected group (n=376,519, mean score=4.28). The mean SP scores were significantly lower for participants with SCZ (mean score=4.08, *P*=6.95e-4) and MDD (mean score=4.22, *P*=4.16e-18) compared to the unaffected group (mean score=4.28; Supplementary Figure 1A; Supplementary Table 1). Mean OF scores across all psychiatric disorders were significantly lower (SCZ mean score=0.001, *P*=3.38e-57; BPD mean score=0.15, *P*=3.53e-33; MDD mean score=0.36, *P*=1.41e-151) compared to the unaffected group (mean score=0.63; Supplementary Figure 1B).The proportion of NEET status assignment in all psychiatric groups also significantly differed from the unaffected group (SCZ *P*=9.65e-259; BPD *P*=1.01e-156; MDD *P*=0; Supplementary Figure 1C; Supplementary Table 1).

### GWAS results

In total, 839 SNPs surpassed the GWS threshold for SP (*P*<5e-8; Manhattan plot in Figure 1A, Q-Q plot in Supplementary Figure 2A, Supplementary Table 2), with 18 lead-SNPs present at 17 independent genomic risk loci (Table 1). LDSC-based SNP-heritability (*h*^2^_SNP_) for SP was 4.1% (S.E.=0.002). For OF, 65 SNPs surpassed the GWS threshold (Supplementary Table 3), with 3 lead-SNPs identified at a single independent locus (chr 8:143,311,088-145,746,556; Figure 1B, Supplementary Figure 2B, Supplementary Table 4). There were no GWS SNPs identified for NEET status (Figure 1C, Supplementary Figure 2C). The *h*^2^_SNP_ for OF was 1.8% (S.E.=0.002) and for NEET status was 1.2% (S.E.=0.001).

**Figure 1:**
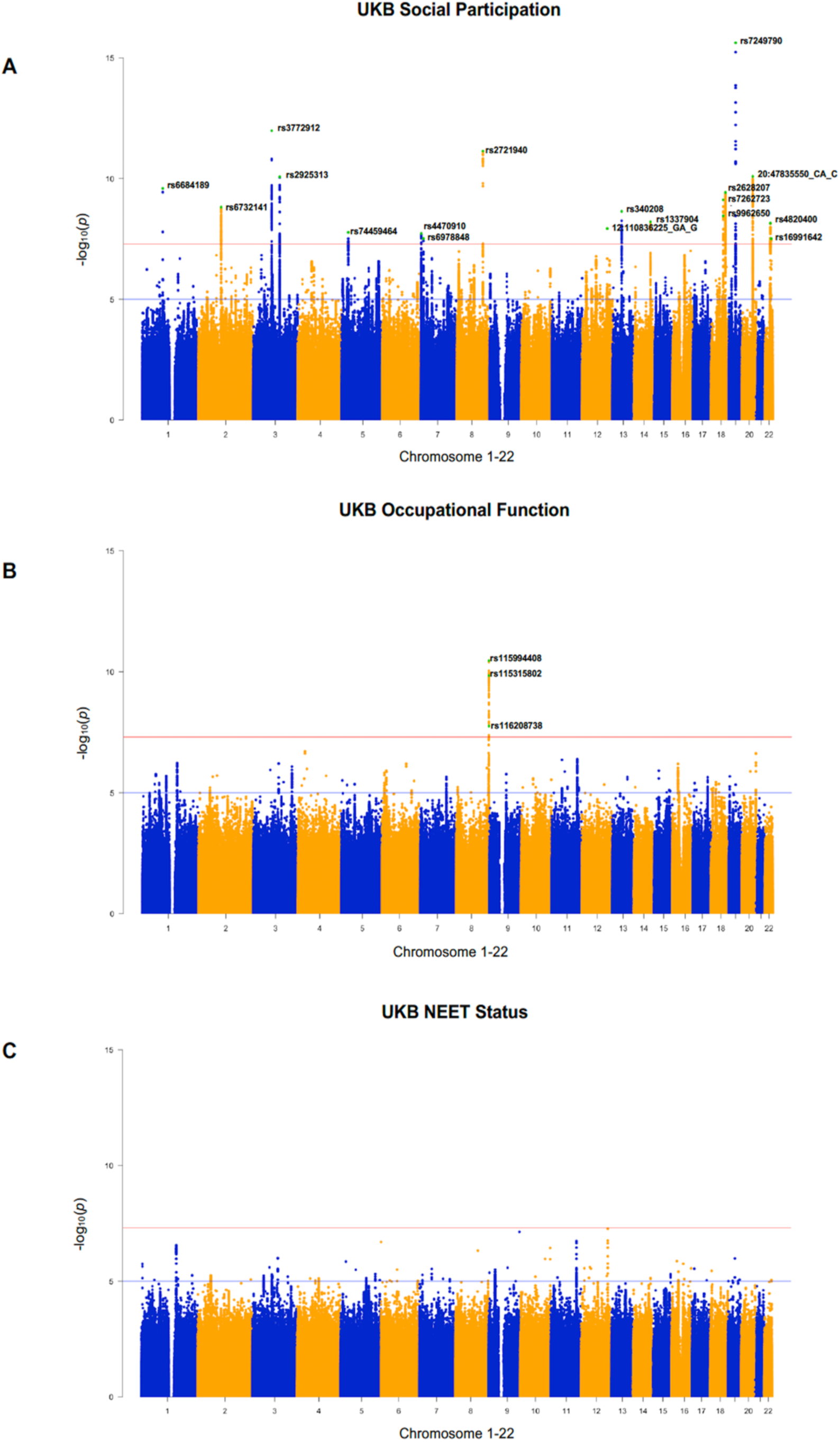
**A** Manhattan plot of the observed -log_10_ *P*-values (*y*-axis) and the distribution of SNPs across chromosomes (*x*-axis) associated with the derived SP variable in the UK Biobank cohort (*n*=404,403). The red line indicates the GWS threshold (*P*<5e-08). Green dots and rsIDs denote the lead variants identified. **B** Manhattan plot of the observed -log_10_ *P*-values of the single nucleotide polymorphisms associated with the derived OF variable in the UK Biobank cohort (*n*=404,569). **C** Manhattan plot of the observed -log_10_ *P*-values of the single nucleotide polymorphisms associated with the derived NEET status variable in the UK Biobank cohort (*n*=404,469).

**Table 1.** Location and significance values of the lead-SNPs at the GWS loci for SP.

| SNP | CHR | POS | A1 | A2 | BETA | P | LOCUS | START<sup>A</sup> | END<sup>A</sup> | GENES<sup>B</sup> |
| --- | --- | --- | --- | --- | --- | --- | --- | --- | --- | --- |
| rs6684189 | 1 | 91093014 | T | C | -0.0209775 | 2.5464E-10 | 1 | 91088884 | 91097495 | U3 |
| rs6732141 | 2 | 100743492 | C | G | 0.0193749 | 1.51525E-09 | 2 | 100602518 | 100835734 | AFF3 |
| rs3772912 | 3 | 81698481 | G | A | 0.0237002 | 1.03979E-12 | 3 | 81499039 | 81986136 | GBE1 |
| rs2925313 | 3 | 117675543 | T | G | 0.0212319 | 8.3773E-11 | 4 | 117493729 | 117821879 | LSAMP |
| rs74459464 | 5 | 29683773 | C | A | -0.0495244 | 1.68067E-08 | 5 | 29596216 | 29713841 | UBL5P1 |
| rs4470910 | 7 | 2071723 | T | C | 0.0234068 | 1.88236E-08 | 6 | 1899447 | 2110850 | MAD1L1, AC069288.1,<br>AC110781.3 |
| rs6978848 | 7 | 11247142 | A | G | 0.0179937 | 3.04755E-08 | 7 | 11244621 | 11253967 | AC004538.3 |
| rs2721940 | 8 | 116636281 | C | A | -0.0224124 | 7.17991E-12 | 8 | 116563879 | 116645056 | TRPS1 |
| 12:110836225_GA_G | 12 | 110836225 | G | GA | 0.019815 | 1.18418E-08 | 9 | 110549533 | 111116655 | ANAPC7, IFT81, VPS29, ARPC3,<br>GPN3, RAD9B, PPTC7, TCTN1,<br>HCVN1, ATP2A2, FAM216A |
| rs340208 | 13 | 60478605 | T | A | 0.0207035 | 2.267E-09 | 10 | 60367014 | 60785511 | DIAPH3 |
| rs13379045 | 14 | 91528977 | C | A | 0.0200233 | 6.10752E-09 | 11 | 91445077 | 91555547 | RPS6KA5, DGLUCY |
| rs72627231 | 18 | 53195964 | T | C | -0.021038 | 7.66951E-10 | 12 | 53195249 | 53621933 | TCF4 |
| rs9962650 | 18 | 53574043 | G | C | 0.0191538 | 3.44831E-09 | 12 | 53195249 | 53621933 | RP11214L13.1 |
| rs2628207 | 18 | 63546386 | T | C | 0.0219922 | 3.78661E-10 | 13 | 63407300 | 63616633 | CDH7 |
| rs7249790 | 19 | 30937444 | G | C | 0.0268073 | 2.39784E-16 | 14 | 30905494 | 30970357 | ZNF536 |
| 20:47835550_CA_C | 20 | 47835550 | C | CA | 0.0219482 | 8.13538E-11 | 15 | 47508077 | 47935069 | DDX27, ARFGEF2, CSE1L,<br>STAU1, ZNFX1, KCNB1 |
| rs4820400 | 22 | 40548084 | T | G | 0.0188991 | 7.02231E-09 | 16 | 40544337 | 40720963 | TNRC6B |
| rs16991642 | 22 | 44595839 | C | T | -0.0191154 | 3.0962E-08 | 17 | 44582264 | 44607514 | PARVG |

<sup>A</sup> START and END defines the locus within which the lead SNPs are located.
<sup>B</sup> GENES includes those mapped to locus co-ordinates. Where no gene mapped to locus co-ordinates, the nearest gene according to ANNOVAR annotations (Ensembl gene build 85) was included. Genes mapped to the associated SNPs by gene prioritisation strategies are also included.

We performed sex-specific analysis to investigate gender-based effects. In total, 33 variants were GWS with 8 lead-SNPs at 8 independent genomic loci in the female-only GWAS of SP (Supplementary Figures 3A and 4A, Supplementary Tables 5 and 6). Six of these eight loci were previously identified in the overall SP GWAS. Although the two remaining novel loci did not reach GWS in the overall GWAS, both lead variants were nominally significant (rs2364959, *P*=0.003; rs4697869, *P*=1.53e-04), suggesting that this is not evidence of a sex-specific effect. No SNP surpassed genome-wide significance in the male-only SP analysis (Supplementary Figures 3B and 4B). For OF, the female-only GWAS revealed 1 significant hit on chromosome 8 (rs560816839), the same locus identified in the overall OF GWAS (Supplementary Figures 5A and 6A). No variant surpassed the significance threshold in the male-only analysis (Supplementary Figures 5B and 6B). No genome-wide significant sex-specific

SNP-based findings were observed for NEET status (Supplementary Figures 7 and 8).

We performed a GWAS of the unaffected group excluding individuals with SCZ, BPD and MDD. For SP, 508 SNPs surpassed genome-wide significance, with 17 lead-SNPs across 16 genomic risk loci (Supplementary Figures 9A and 10A; Supplementary Tables 7 and 8). Just one new locus on chromosome 12 was identified (12:61097046_TC_T). While this locus did not reach GWS in the overall SP GWAS, it did achieve nominal significance (*P*=1.66e-07). Overall, the beta values for all SNPs from both analyses were highly correlated (R^2^_adjusted_=0.89, *P*<2.2e-16). There were 17 genome-wide significant SNPs for OF, with 3 lead-SNPs at a single genomic locus previously identified in the overall GWAS (Supplementary Figure 9B & 10B, Supplementary Table 9 & 10) and once again, no genome-wide significant hits for NEET status (Supplementary Figure 9C & 10C). Overall, these data indicated that inclusion of individuals with a diagnosis of a psychiatric disorder was not a major driver of the genetic associations detected for SP or OF.

### Statistical fine-mapping

Using the Bayesian fine-mapping approach, SuSiE, we identified fifteen 95% credible sets of potentially causal variants for 13 genomic loci associated with SP (Supplementary Table 11). Overall, 3 credible causal variants (CCVs) with a PIP score >0.5 were identified, each representing an independent association signal at a different genomic risk locus. All three CCVs were previously defined as the lead variant at all three genomic regions (rs3772912 (chr 3, intron of *GBE1*), PIP=0.76; rs7249790 (chr 19, intron of *ZNF536*), PIP=0.70; rs6684189 (chr 1, upstream of *U3*), PIP=0.70). Although variants within the subsequent 95% credible sets did not exceed the PIP score threshold (>0.5), the lead SNP at 8 of these genomic loci acquired the highest PIP and purity score. Variants with the greatest PIP score were not the lead SNPs at two genomic risk loci (Supplementary Table 11). No 95% credible sets of potentially causal variants were identified for the one OF locus.

### Functional Annotation

CADD scores >12.37 (i.e. minimum threshold for pathogenicity) were observed for 77 SP-associated SNPs (3.7%; Supplementary Table 12) across 15 of the significant genomic risk loci. Four of the six SNPs with the highest CADD scores (>20) were missense variants (rs1130146 and rs11553387 in *DDX27*, rs2229519 in *GBE1* and rs2291343 in *CDH7*). Nineteen SNPs (3.4%) exceeded the CADD score threshold for OF (Supplementary Table 13) at the single significant genomic locus and included rs114379623 (a missense variant in *TOP1MT*) and rs115510595 (a 3’UTR variant in *ZNF696*).

### Gene-based analysis

MAGMA gene-based analyses identified 17 genes that surpassed the significance threshold (*P*<2.63e-6) for the SP phenotype (Supplementary Table 14) and the top genes included *CDH7* (*P*=5.57e-12), *GBE1* (*P*=5.90e-11) and *ZNF536* (*P*=1.20e-10). For OF, 3 genes including *DRD2* (*P*=4.47e-08), *TNRC6A* (*P*=1.75e-06) and *TTC12* (*P*=2.03e-06) surpassed the significance threshold (Supplementary Table 15), while *DRD2* (*P*=2.77e-07) was the single gene was identified for NEET status (Supplementary Table 16).

### Gene-set and tissue analysis

Investigating biological functions, gene-set analysis showed that the gene-set responsible for the *regulation of synaptic structure/activity* was significantly enriched for genes associated with the SP phenotype (P_bon_=0.03). No gene-set passed Bonferroni correction for either OF or NEET status phenotypes.

Based on 53 tissue types from GTEx v8, the MAGMA analysis revealed that SP-associated genes were most significantly enriched across 11 brain regions including the cerebellum, cortex, hippocampus, basal ganglia, hypothalamus, and amygdala (Supplementary Figure 11). Overall, these genes were generally implicated in brain and pituitary tissues (Supplementary Figure 12). The genes associated with OF and NEET status were not significantly enriched in any specific tissue-type (Supplementary Figures 13-16).

### Gene Mapping

Seventy-eight unique genes were implicated for SP (30 genes by positional gene-mapping, 25 genes by eQTL-mapping and 57 genes by 3D chromatin-mapping). Of the 17 genes identified by the earlier gene-based MAGMA analysis, 12 overlapped with those identified by the gene-mapping approaches. In total, three genes were observed across all four approaches: *CSE1L*, *STAU1*, and *TNRC6B* (Figure 2A; Supplementary Table 17).

**Figure 2:**
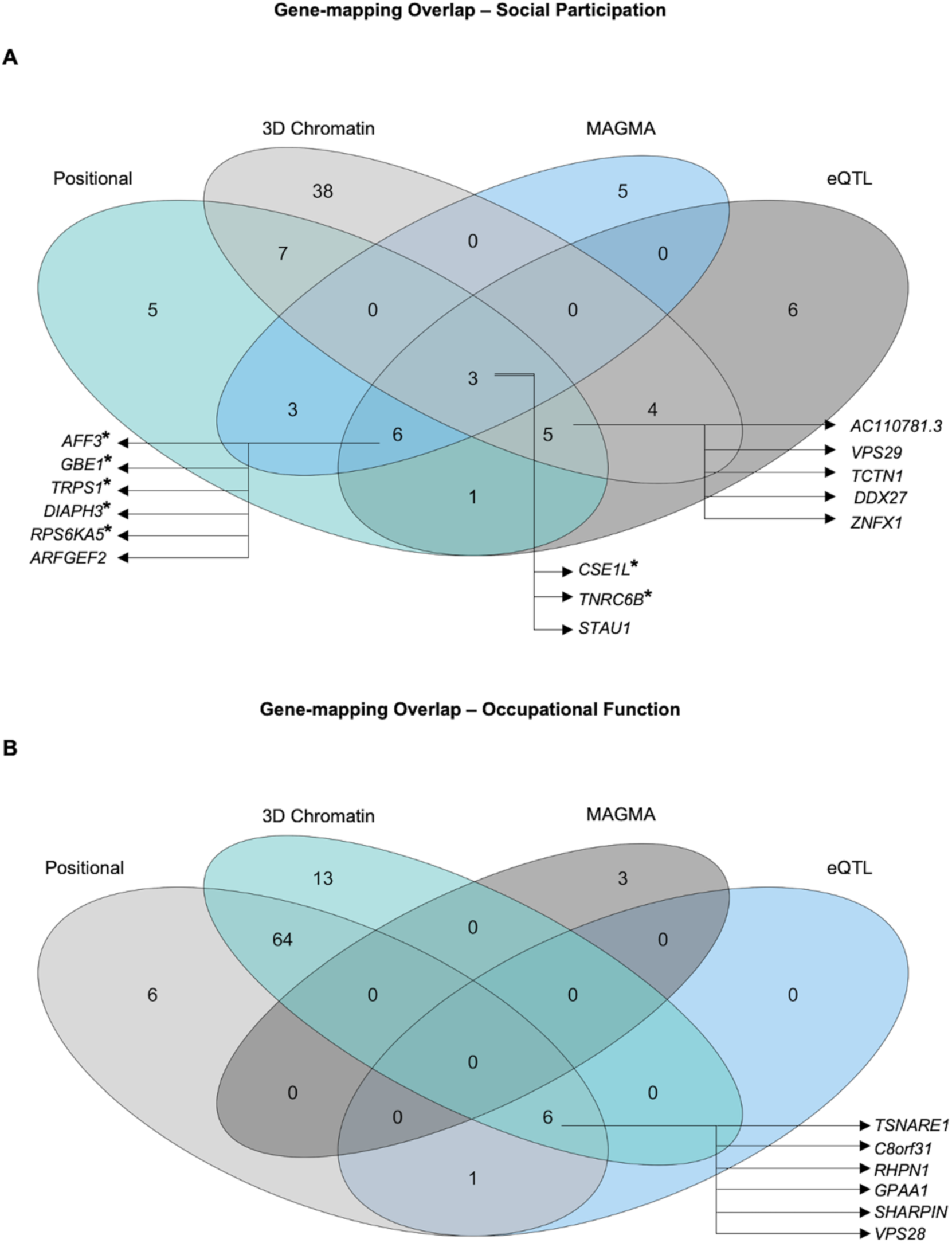
Venn diagram visualising the number of genes mapped to the **A** SP and **B** OF phenotypes by each approach and the number of overlapping genes between outputs. Asterix (*) denotes the genes also prioritized by PoPS.

Seventy-seven individual genes were positionally mapped for OF. Subsequent eQTL and 3D chromatin interaction mapping provided support for 7 genes and 83 genes respectively. Across approaches, 65 genes were identified by at least two approaches and 6 genes were implicated by all strategies (*C8orf31*, *GPAA1*, *RHPN1*, *SHARPIN*, *TSNARE1*, and *VPS28*) although none of these genes overlapped with those identified by the gene-based MAGMA analysis (Figure 2B; Supplementary Table 18).

All 47 genes implicated in NEET status were positional as the absence of genome-wide significant symptoms prevented further analysis. This included the single gene identified by MAGMA analysis, *DRD2*, for NEET status (Supplementary Table 19).

### Gene Prioritization

We computed PoPS scores for all protein coding genes within 500-kb of the 869 and 65 GWS associations for SP and OF, respectively (PoPS was not performed for NEET status as no variant exceeded the GWS threshold). This resulted in the prioritization of 16 genes for SP (Supplementary Table 20) and 2 genes for OF. The highest scoring genes for SP according to PoPS were *CDH7*, *GBE1*, and *ZNF536*, the same top three genes implicated by the MAGMA analysis. Of the 16 genes prioritized for SP, two genes, *TNRC6B* and *CSE1L*, were previously implicated across all three gene-mapping approaches (Supplementary Table 21). For OF, both prioritized genes, *LY6H* and *PARP10*, were previous mapped using two approaches, positional and 3D chromatin interaction mapping.

### Transcriptome-wide association analysis

A transcriptome wide association study (TWAS) was performed for SP and OF using FUSION and eQTL gene expression data from the PsychENCODE Consortium.^49^ Of the 11,934 genes analysed, the expression of 6 genes encompassing 4 independent genomic risk loci were significantly associated with SP (*P*-Bon<4.19e-06; Supplementary Figure 17A; Supplementary Table 22). Among the significant genes were *TRSP1* (*P*=2.08e-12), *CSE1L* (*P*=3.95e-09), and *STAU1* (*P*=7.22e-07), all of which were previously mapped to the SP phenotype. No genes were identified for OF (Supplementary Figure 17B). TWAS fine mapping was performed to prioritize the most likely causal gene in each region for SP. Within the 90%-credible set, FOCUS prioritized 5 genes across 5 distinct genomic loci, with a PIP score >0.8. This included *TRPS1*, *CSE1L*, *URI1*, *AFF3*, and *GBE1*

(Supplementary Table 23).

### Genetic correlations

LDSC analysis was performed to investigate the genetic correlations between SP, OF and NEET status and psychiatric disorders of interest (Figure 3). Significant genetic correlations were found between lower SP and greater risk of ADHD, ASD, SCZ, and MDD but not BD. Greater risk of ADHD, BD, MDD, and SCZ was significantly genetically corelated with both lower OF and being NEET.

**Figure 3:**
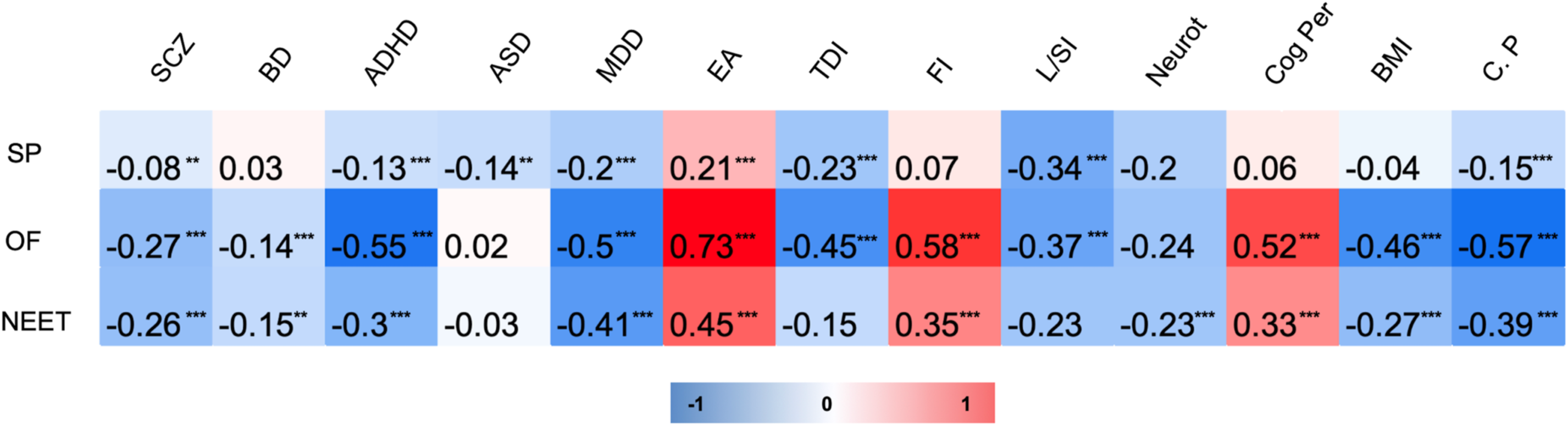
Heatmap of the genetic correlation results from LD score regression and batch genetic correlation for SP, OF, and NEET status with relevant traits of interest. Asterix indicates a significant genetic correlation (** <0.01, ***<0.001; multiple testing threshold *P*<1.39e-5). Colour legend depicts strength of the positive/negative correlations. EA: Educational Attainment; TDI: Townsend Deprivation Index; FI: Fluid Intelligence; L/SI: Loneliness/Social isolation; Neurot: Neuroticism; Cog Per: Cognitive Performance; BMI: Body Mass Index; C. P: Chronic Pain. Note: To aid interpretation, NEET status values have been reversed.

After correction for multiple testing, the batch genetic correlation revealed that SP, OF, and NEET status showed significant genetic correlations with 39, 365, and 140 traits respectively from across 1,583 published GWASs (Supplementary Tables 24, 25 & 26). Lower SP was genetically correlated with greater loneliness and greater socioeconomic deprivation and lower educational attainment (Figure 3). Lower OF was genetically correlated with greater loneliness, and greater socioeconomic deprivation (TDI), while higher OF was genetically correlated with greater educational attainment, higher cognitive performance, and higher fluid intelligence (Figure 3). NEET status was genetically correlated with lower educational attainment, lower fluid intelligence, lower cognitive performance and higher scores for neuroticism (Figure 3). Interestingly, greater chronic pain was genetically correlated with lower scores for all three phenotypes.

### Mendelian Randomization

MR was performed to investigate the causal relationships between SP and psychiatric disorders (SCZ, BD, MDD, ADHD, and ASD) and EA. SP did have a nominally significant causal effect on MDD while both BD and ADHD had nominally significant causal effects on SP. For the occupational engagement phenotypes, ADHD had a significant causal effect on OF (*P*=0.00037) and SCZ had a significant causal effect on NEET (*P*=0.00061; Figure 4; Supplementary Table 27) with both results surviving multiple test correction. This indicates vertical pleiotropy between these two disorders and our UKB occupational engagement phenotypes.

**Figure 4:**
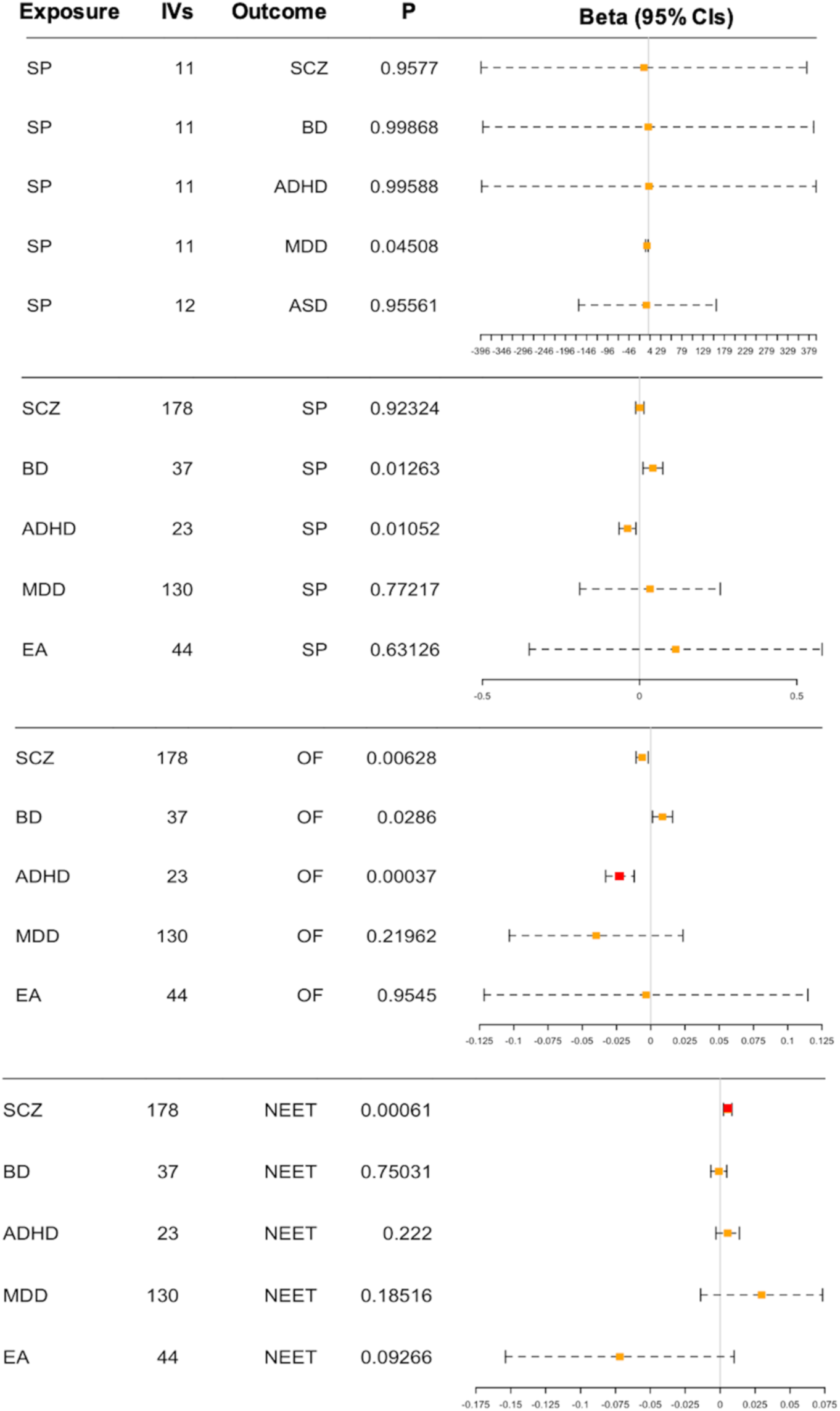
MR forest plot of exposure (SP, SCZ, BD, ADHD, MDD, EA) versus outcomes (SCZ, BD, ADHD, MDD, ASD, SP, OF, NEET status). Plot shows the 95% confidence intervals (CIs) of the effect estimates (β). Each line represents the CI and red dots identify associations that reach the *P*_bon_ threshold.

## Discussion

To our knowledge, this is the first study to investigate the genetics of objective social functioning using UKB data, which included aspects of leisure/social activity and frequency of visits with friends/family to create the SP phenotype. Here, we define objective functioning as a measure of active participation and engagement, capturing how well an individual outwardly functions despite subjective emotional influence. Although active functioning has been previously incorporated into social phenotypes,^22,24,25^ our aim was to solely capture outward functioning. We also investigated the genetics of occupational engagement, that is, the extent to which an individual actively engages occupationally/vocationally.

We observed that mean SP scores were significantly lower for participants with SCZ and MDD in the UKB. Mean OF scores were significantly lower and the proportion of individuals that were NEET was significantly higher across all psychiatric disorders tested. This indicated that our phenotypes indexing psychosocial disability were relevant to psychotic and other psychiatric disorders. By performing GWASs of these phenotypes excluding individuals with psychiatric disorders for comparison with our overall GWASs, we demonstrated that inclusion of these individuals is not responsible for the genetic associations detected. We detected no major sex-specific effects in our association analysis.

### Genetics of SP

We interpreted the GWAS results using a series of gene mapping approaches (positional, eQTL and 3D chromatin-mapping) in combination with the MAGMA gene-based association test, gene prioritization (using PoPs)^47^ and TWAS. *CSE1L* was identified by all methods. It plays a role in cell proliferation and apoptosis^26^ and is a potential target gene for miR-137, a well-established SCZ risk locus.^60^ Increased expression of *CSE1L* has been associated with an elevated risk for SCZ.^61^ The *STAU1* gene, identified by all methods except gene prioritization, maps to the same locus as *CSE1L*. This is a multifunctional double-stranded RNA-binding protein involved in neuronal differentiation, often overexpressed in individuals with neurodegenerative diagnoses.^62^ *TNRC6B* was also consistently associated with SP by all strategies, except TWAS. It is involved in translational inhibition,^63^ which has been repeatedly linked to intellectual disability, ASD and ADHD traits, and subsequent behavioural abnormalities.^64^

The strongest association signal for SP was at *ZNF536* where the lead SNP was identified by SuSiE fine-mapping^43^ as a credible causal variant. This is a transcriptional repressor gene involved in regulating neuronal differentiation.^65^ *ZNF536* was previously implicated in a recent GWAS of social-isolation.^25^ There is also evidence that *ZNF536* increases risk for neurodevelopmental disorders characterized by social deficits, including SCZ and ASD.^66^ Rare truncating variants at *ZNF536* have been observed in individuals with ASD.^67^

The second strongest association was at *GBE1* where the associated haplotype contains a missense variant in exon 5 of the gene. *GBE1*, involved in glycogen production, was previously implicated in GWASs of sociability.^24^ The lead variant at *GBE1* has also been implicated in neuroticism,^68^ a trait often accompanied by avoidance of social situations due to social unease.^69^ The associated haplotype at two additional genes contained also a missense variant; *CDH7* and *DDX27*. *CDH7*, a calcium dependent protein coding gene, had the highest gene prioritization score. Variants at this gene have been previously associated with neuroticism,^70^ and cognitive function,^30^ factors known to influence SP. *DDX27* is an RNA helicase implicated in altering RNA secondary structure and is commonly linked to muscle growth^71^ as well as various cancer types.^72,73^ It is worth noting that *DDX27* maps to the same locus as *CSE1L* and *STAU1*.

There was evidence from our GO analysis to suggest that pathways involved in the regulation of synaptic signalling may play a role in the biology of SP. Interestingly, enrichment of genes in synaptic pathways has been shown in multiple psychotic disorders, including SCZ^26,52^ and BD.^53^ Further probing into the relationship between SP-associated genes and neuropsychiatric disorders (SCZ, BD, MDD) revealed a notable link between SCZ and *TCF4*.^52^ *TCF4* is a transcription factor gene with a substantial role in neural development^74^ repeatedly associated with neurodevelopmental disorders including SCZ.^75,76^ Our lead SP variant at *TCF4* (rs72627231) both surpassed GWS in the most recent SCZ GWAS^52^ and is in high LD (*r^2^*=0.52) with one of the lead SCZ variants at that same locus, suggesting independent association of the same variant at *TCF4* with both phenotypes.

### Genetics of OF and NEET

The current work is among the few studies that have investigated the genetic basis of occupational engagement, as defined by occupational status. Here, our only GWS locus for OF contained three lead SNPs, each of which had a low minor allele frequency. Gene mapping approaches proposed six genes in the region (*C8orf31*, *GPAA1*, *RHPN1*, *SHARPIN*, *TSNARE1*, and *VPS28*) and PoPs gene prioritization proposed another two (*LY6H* and *PARP10*). Functional annotation indicated that one of the associated haplotypes included a missense variant in *TOP1MT*. It is thus a difficult region to tease apart. *TSNARE1*, strongly associated with SCZ,^77^ belongs to a group of proteins believed to increase SCZ risk via synaptic dysfunction.^78^ *LY6H*, an acetylcholine receptor binding gene, is putatively involved in neurodegeneration in Alzheimer’s disease.^79^ *PARP10* is implicated in the regulation of gene transcription and plays a undefined role in the pathogenesis of ASD.^80^

The MAGMA gene-based association test identified three GWS genes for OF, but these were not located at the single GWS significant locus for OF on chromosome 8. These genes are *DRD2* and *TTC12* on chromosome 11, and *TNRC6A* on chromosome 16. Here, individual SNPs approached GWS levels such that in combination within the gene-based test, they produced a GWS result at the gene level. It was similar for NEET where there were no GWS SNPs but one GWS gene, *DRD2*. *DRD2* encodes the dopamine D2 receptor and is a notable target of most effective antipsychotic drugs and is associated with SCZ.^52^ The most associated SNPs for OF (rs7125588) and NEET (rs4309187) are not in LD with each other, but each is in high LD with one of the two independent lead SNPs for SCZ. This suggests independent association of variants at *DRD2* with OF and SCZ and with NEET and SCZ.

### Correlations and causal relationships

Genetic correlations showed evidence of a shared genetic etiology between our phenotypes and major psychiatric disorders, personality traits, cognitive phenotypes, educational attainment, and measures of loneliness, social isolation, social deprivation and other health outcomes, e.g. BMI and chronic pain. These results suggest that SP, OF and NEET cut across both mental and physical health generally. We showed a negative correlation between SP and neuroticism, lending support to the previously established link between social impairment and anxiety^24^ and the idea that there may be common genetic factors driving reduced social functioning and increased neuroticism.^25^

Despite SNP-based heritability estimates being low at 1-4% for SP, OF and NEET, the MR analysis revealed evidence of causality of psychiatric disorders on these phenotypes; ADHD being causal for lower OF and SCZ being causal for an increased likelihood of having NEET status. In reverse, there was some evidence that reduced SP may be causal for MDD. Further evidence for this causal link is required but if established, it supports targeting SP by intervention to reduce the burden of MDD in the population.

### Strengths and limitations

A major strength of the study is the large sample size made possible by using fastGWA,^36^ a GWAS method which allowed us maximize our usable sample by retaining related individuals, increasing the power of our study. Another strength is that we used active functioning measures to avoid the influence of individual subjective feelings. This, however, also proved to be one of the main limitations, in that, trying to measure active functioning limited suitable UKB social and occupational variables. Data is limited in the UKB due to the stringent multiple-choice/Likert scales implemented within the ACE. Another limitation is that participants in this sample were of white British ancestry, meaning our results are not generalizable to other ancestral groups. It should also be noted that all data utilized within the current study was cross-sectional and from a single data collection timepoint. Future research should aim to incorporate longitudinal measures to study the underlying genetics of active social and occupational functioning. Finally, it was not possible to extend the MR analysis of causal relationships between SP, OF and NEET and several measures because the genetic studies of those phenotypes have used the UKB and thus, genetic data from large independent samples was not attainable.

## Conclusion

We have demonstrated a significant genetic component to PD by generating and then investigating our SP and OF phenotypes. Using a combination of gene mapping, prioritization and association tests, we have established a list of genes influencing these phenotypes including some also associated with SCZ. The low heritabilities detected may indicate that other genetic factors not yet identified are at play here. This may alternatively indicate that environmental factors account for much of the individual variation in psychosocial functioning in the population. However, despite this, there is evidence of a shared aetiology between PD and neuropsychiatric disorders and other psychological and health-related phenotypes. In addition, there is evidence for small but significant causal relationships between neuropsychiatric disorders and occupational engagement that require further exploration.

## Data Availability

All data produced in the present study are available upon reasonable request to the authors

