## Supplementary Methods for "Genome-wide association studies of social participation and occupational engagement in the UK Biobank"

Population

All 502,292 UKB participants were registered with the National Health Service and resided ~25 miles from one of the 23 centres. Participants for the current study were those who took part in initial baseline assessment and answered questions on leisure activity, frequency of visits with friends and family, and current employment status. Genotype data was available for 487,409 individuals. Filtering of these individuals was performed using available UKB data on white-British ancestry (Data-Field 22006), discordant sex, sex aneuploidies, missing phenotype, withdrawn consent, and covariate data. The sample size for each GWAS was dependent on the availability of phenotypic and genotypic data.

Phenotypes

Participants completed a self-report, touchscreen questionnaire using the Assessment Centre Environment (ACE), a direct data entry system. Using baseline (i0) UKB data from each participant, we derived three unique variables: social participation (SP), occupational function (OF), and not in education, employment, and training (NEET) status.

We defined SP as the degree to which an individual participates in social activities and/or social interactions. This is a measure derived using variables representative of objective social function (i.e. how a person *actively* participates in society) and not subjective function (i.e. how a person views their participation in society). Leisure/social activities was determined by the question “Which of the following do you attend once a week or more often?”. The following choices were provided 1= “Sports club or gym”; 2 = “Pub or social club”; 3 = “Religious group”; 4 = “Adult education class”; 5 = “Other group activity”; -7 = “None of the above” or -3 = “Prefer not to answer”, with the option to select more than one. Frequency of friends/family visits was assessed with the question “How often do you visit friends or family or have them visit you?” and scored with 1 = “Almost daily”; 2 = “2-4 times a week”; 3 = “About once a week”; 4 = “About once a month”; 5 = “Once every few months”; 6 = “Never or almost never”; 7 = “No friends/family outside of the household”; -1 = “Do not know” or -3 = “Prefer not to answer”.

To derive the composite SP score, both variables were recoded. For leisure/social activities, participants were scored by summing the number of activities selected (i.e. “Pub or social club” + “Religious Group” + “Other group activity” = 3).

Frequency of friends/family visits was reverse coded whereby a higher score indicated greater levels of SP (5 = “Almost daily; 0 = “Never or almost never”). Those who answered “No friends/family outside of the household”, “Do not know”, “Prefer not to say”, or “None of the above” were considered “NA”. To calculate SP, the new scores from both variables were simply summed producing a composite value where higher scoring indicated greater SP.

OE was defined as the degree to which an individual engages in occupational/vocational activities. Both variables, OF and NEET status, were derived from UKB variable, current employment status (Data-Field 6142). NEET is a term which originated in youth mental health research relating to the transition from education into work and was coined in a 1999 UK report called “Bridging the Gap”.^1^ Current employment status was determined by the question “Which of the following describes your current situation?”. The choices provided included 1 = “In paid employment or self-employed”; 2 = “Retired”; 3 = “Looking after the home and/or family”; 4 = “Unable to work because of sickness or disability”; 5 = “Unemployed”; 6 = “Doing unpaid or voluntary work”; 7 = “Full or part-time student”; -7 = “None of the above” or -3 “Prefer not to answer”. Participants could select more than one answer.

For OF, answers to the current employment variable were recoded into 1 = “Good” (In paid employment or self-employed, doing unpaid or voluntary work, full-time or part-time student, and looking after the home and/or family); 0 = “Neutral” (Retired) or -1 = “Bad” (Unemployed and unable to work because of sickness or disability). Those who selected “None of the above”, “Prefer not to answer” or who provided conflicting answers (i.e. selected employed and unemployed) were deemed “NA”. A composite score was derived by simply summing the recoded scores (i.e. “looking after the home and/or family” = 1 + “doing unpaid or voluntary work” =1 + “unemployed” = -1: composite score = 1).

NEET status was coded into a binary variable of 0 = “non-NEET status” and 1 = “NEET status” and then later recoded into 1 = “non-NEET status” and 2 = “NEET status” for the purpose of analysis. Individuals who were not in education, employment, or training were deemed to be of NEET status (alone or any combination of the following: doing unpaid or voluntary work, looking after the home, retired, unemployed, and/or unable to work because of sickness or disability).

Phenotypic Data on Disorders of Interest

The ICD-10 diagnosis codes used for this exploratory analysis were as follows; SCZ (F20-F29), BD (F31), MDD (F32-F39), and ADHD (F90). As data was not normally distributed, the Poisson general linear model was used for SP and OF, while the binomial model was used for NEET status. As OF included negative scoring (ranging from -2 to +4), +2 was added to all participant OF scores to create non-negative integers, facilitating the analysis.

Genetic Data

The Applied Biosystems UK Biobank Axiom Array^2^ was used for 363,719 samples in the present study, while the Applied Biosystems UK BiLEVE Axiom Array^3^ was used for 44,562 samples.

Statistical Analyses

Genotype analyses were performed using JupyterLab on the UKB Research Analysis Platform (RAP; https://ukbiobank.dnanexus.com), a cloud-based resource. The analyses tested the effect of 13,123,036, 13,122,864, and 13,122,845 SNPs with a MAF>0.01 for SP, OF, and NEET status respectively.

FastGWA is designed to simultaneously account for relatedness and other population stratification through generation of a genetic relationship matrix (GRM),^4^ making it a resource-efficient method for large scale datasets like the UKB. Inclusion of a GRM maximises the sample by retaining related individuals rather than removing them, as prescribed by the more established approaches (e.g., PLINK). A sparse GRM was generated for individuals of white-British ancestry in 250 consecutive partitions using UKB genotype data, which had been pruned using HapMap3 SNPs (https://alkesgroup.broadinstitute.org/LDSCORE/w_hm3.snplist.bz2) (window size of 100-kilobases, step size of 10, and a linkage disequilibrium (LD) *r^2^* threshold of 0.01). Pruned SNPs with a MAF>0.01 were retained for each chromosome and then merged into a single set of p-files. PLINK (v2.0) (www.cog-genomics.org/plink/2.0/) was used to omit multiallelic SNPs and existing p-files were converted to b-file format for creation of the GRM, as recommended. This process reduced the number of SNPs comprising the sparse GRM to 68,273, rather than the available ~15 million. Utilisation of the sparse GRM increased our overall sample size by ~27,000.

FastGWA was also used to conduct the GWASs of the three phenotypes within sex specific samples (male vs female) and a separate unaffected group to explore the influence of genetic sex or a neuropsychiatric diagnosis on the overall results. To test the influence of genetic sex on the UKB phenotypes, GWASs were performed on variants for female (SP=13,121,634; OF=13,121,259; NEET status=13,121,211) and male (SP=13,116,985; OF=13,116,858; NEET status=13,116,838) participants independently (MAF>0.01). The sample was divided into males and females using the data from the UKB sex variable (Data Field 31). All parameters remained the same for the genome-wide analysis.

An exploratory GWAS was carried out testing the effect of 13,122,336, 13,122,374, and 13,122,279 variants for SP, OF, and NEET status respectively using the unaffected group (MAF>0.01). The unaffected group excluded individuals with an ICD-10 neuropsychiatric diagnosis (SCZ, BPD, MDD, and ADHD; *n*=35,859), who were removed during creation of the GRM and from the final fastGWA. All other parameters remained the same and both analyses were adjusted for age, sex (unaffected group only), array, TDI scores, assessment centre and the first 20 principal components.

Fine-mapping

Using PLINK2 (v2.0) (www.cog-genomics.org/plink/2.0/), the European panel of the 1000 Genomes Project (Phase 3)^5^ (https://vu.data.surfsara.nl/index.php/s/VZNByNwpD8qqINe) was harmonised with the Genome Reference Consortium Human Build 37 (GRCh37)^6^ data to maintain consistency across reference alleles. The harmonised European panel data was then subsequently used as the LD reference. Duplicate SNPs were removed from fastGWA summary statistics, which was then filtered to contain common SNPs existing within both the harmonised and summary statistics datasets. Lead SNPs were isolated from summary statistics using gwaslab^7^ in Python (v 3.9.16) (https://github.com/Cloufield/gwaslab). The 95% credible sets of causal variants were identified for each SNP signal at the known loci for both phenotypes using the susieR package (v 0.12.35)^8^ in R (v4.4.0).^9^ Variants within a window of +/-500-kb either side of a lead SNP signal were defined (R, v4.4.0) and then extracted (PLINK, v1.9)^10^ to create the LD matrix in susieR. Posterior inclusion probability (PIP) scores were assigned to all variants identified within 95% credible sets. All fine-mapping analyses were carried out using Bash, Python, and R within JupyterLab Notebook (v2.5.0) on the UKB RAP.

Gene-mapping

(1) Positional mapping: SNPs were mapped to genes if located within a 10kb window from protein-coding genes within the reference panel

(2) eQTL mapping: mapped SNPs to genes where SNPs were associated with altered expression of a gene within a 1Mb window (i.e. cis-eQTL). This was based on brain expression data from PsychENCODE (http://resource.psychencode.org/),^11^ the CommonMind Consortium (https://www.synapse.org//#!Synapse:syn5585484),^12^ BRAINEAC (http://www.braineac.org/), and GTEx v8 Brain (http://www.gtexportal.org/home/datasets). A false discovery rate (FDR) of <0.05 was used identify significant eQTL SNP-gene pairs

(3) 3D chromatin interaction mapping: SNPs were mapped to genes based on chromatin interactions using PsychENCODE (EP links and Promoter anchored loops; http://resource.psychencode.org/), Hi-C adult cortex and fetal cortex,^16^ and Hi-C dorsolateral prefrontal cortex and hippocampus data (https://www.ncbi.nlm.nih.gov/geo/query/acc.cgi?acc=GSE87112). The default FDR (<1e-6) and promoter region window (250bp upstream and 500bp downstream of the gene’s transcription start site) were implemented. Annotation of the enhancer and promoter regions was carried out using the 12 brain-related epigenomes from the Roadmap Epigenome Project (http://egg2.wustl.edu/roadmap/web_portal/DNase_reg.html#delieation)

Transcriptome-wide association study

The fastGWA summary statistics for SP and OF were formatted using the munge function within FOCUS (v0.09)^13^ as this avoids restricting the analysis to HapMap3 variants. PsychENCODE gene expression weights were downloaded from the PsychENCODE resources page under the section ‘Cross-Disorder Analysis TWAS weights’ (http://resource.psychencode.org/). The genomic position information file was generated using the biomaRt (v 2.58.2)^14^ (https://github.com/Huber-group-EMBL/biomaRt) package in R with the genome assembly GRCh37.^6^ Gene Ensembl IDs were used to obtain gene symbol, chromosome, start position and end position information. Any gene for which all such information was not obtained was removed from analysis. The European panel of the 1000 Genomes Project (Phase 3)^5^ (https://vu.data.surfsara.nl/index.php/s/VZNByNwpD8qqINe) was used as the LD reference panel. Variant IDs of the PsychENCODE SNP-weights were formatted as chromosome and bp position (CHR:BP), rather than RSIDs. To maintain consistency, the RSID columns of the summary statistics and the LD reference panel were converted to the chromosomal coordinate format using R.

FOCUS (v0.09)^13^ was used to perform TWAS fine-mapping. Using JupyterLab on the UKB RAP, FOCUS fine-mapping was carried out using a conda environment as recommended.^13^ The FOCUS ‘import’ function was used to convert the FUSION weight-list genomic position information file (.pos) to the FOCUS-specific sqlite database format (.db). As PsychENCODE Consortium weights were utilised, the prior probability and locations parameters were adjusted to use ”gencode37” and ’37:EUR’ data resources. PIP scores were assigned to each gene and 90% credible gene sets were computed to explain observed genomic risk.

Bi-directional Mendelian Randomisation

To perform the bi-directional mendelian randomisation, LD clumping was performed using the clump_data function from the TwoSampleMR (v0.6.8)^15^ R package on default settings. Any SNP within a 10,000 kb of the top hits were removed to ensure independence and only SNPs within the 1000 Genomes Project European sample^5^ (http://fileserve.mrcieu.ac.uk/ld/1kg.v3.tgz) with *r*^2^ > 0.001 remained. Exposure and outcome datasets were harmonised using TwoSampleMR^15^ function harmonise_data before running the final analysis.
