## Supplementary Figures for "Genome-wide association studies of social participation and occupational engagement in the UK Biobank"

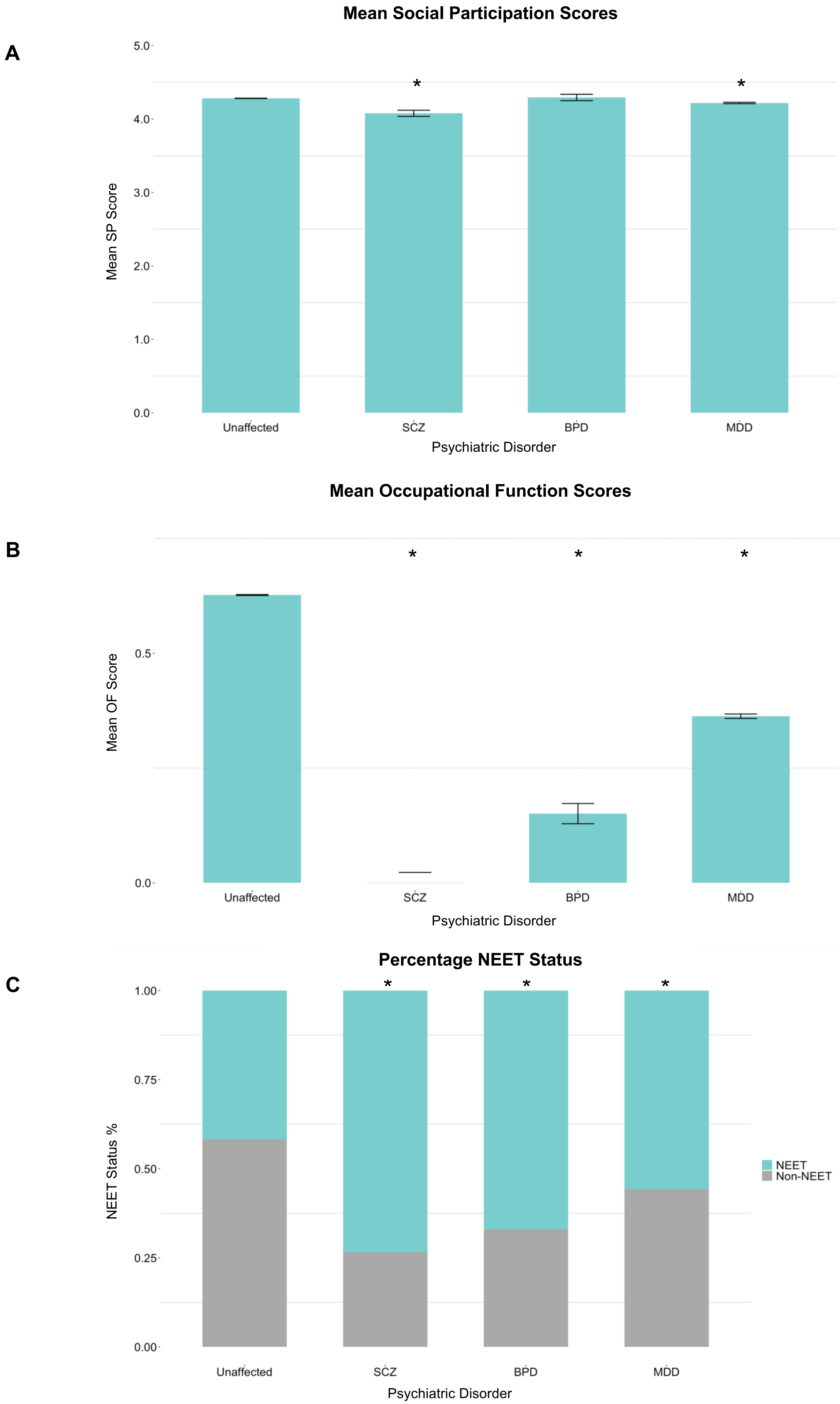
A
Mean Social Participation Scores
Mean SP Score
5.0
4.0
3.0
2.0
1.0
0.0
*
*
Unaffected
SCZ
BPD
MDD
Psychiatric Disorder
B
Mean Occupational Function Scores
Mean OF Score
0.5
0.0
*
*
*
Unaffected
SCZ
BPD
MDD
Psychiatric Disorder
C
Percentage NEET Status
NEET Status %
1.00
0.75
0.50
0.25
0.00
*
*
*
NEET
Non-NEET
Unaffected
SCZ
BPD
MDD
Psychiatric Disorder

**Supplementary Figure 1** Bar charts of mean SP (**A**) and OF (**B**) scores and percentage split in NEET status assignment (**C**) across groups (Unaffected, SCZ, BD, & MDD). Error bars show the 95% confidence intervals (S.E.). An Asterix indicates a significant difference in mean scoring of psychiatric disorder groups compared to the unaffected group (*). **Note**: (1) the y-axis differs according to phenotype, (2) ADHD is reported on in Supplementary Table 1.

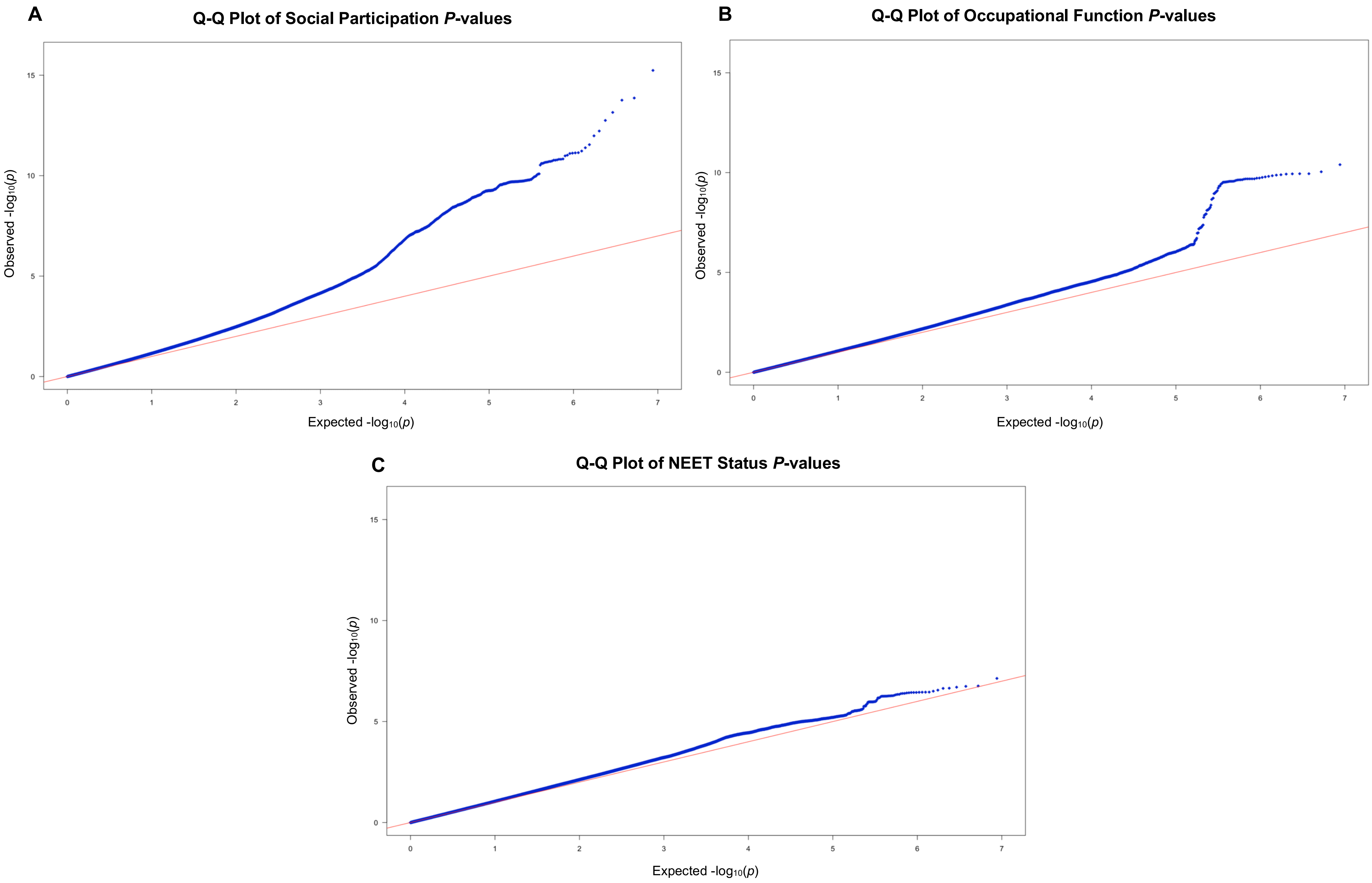


**Supplementary Figure 2** Q-Q plot of *P*-values for the SNP-based association analyses of the UKB phenotypes: **A** SP, **B** OF, and **C** NEET status.

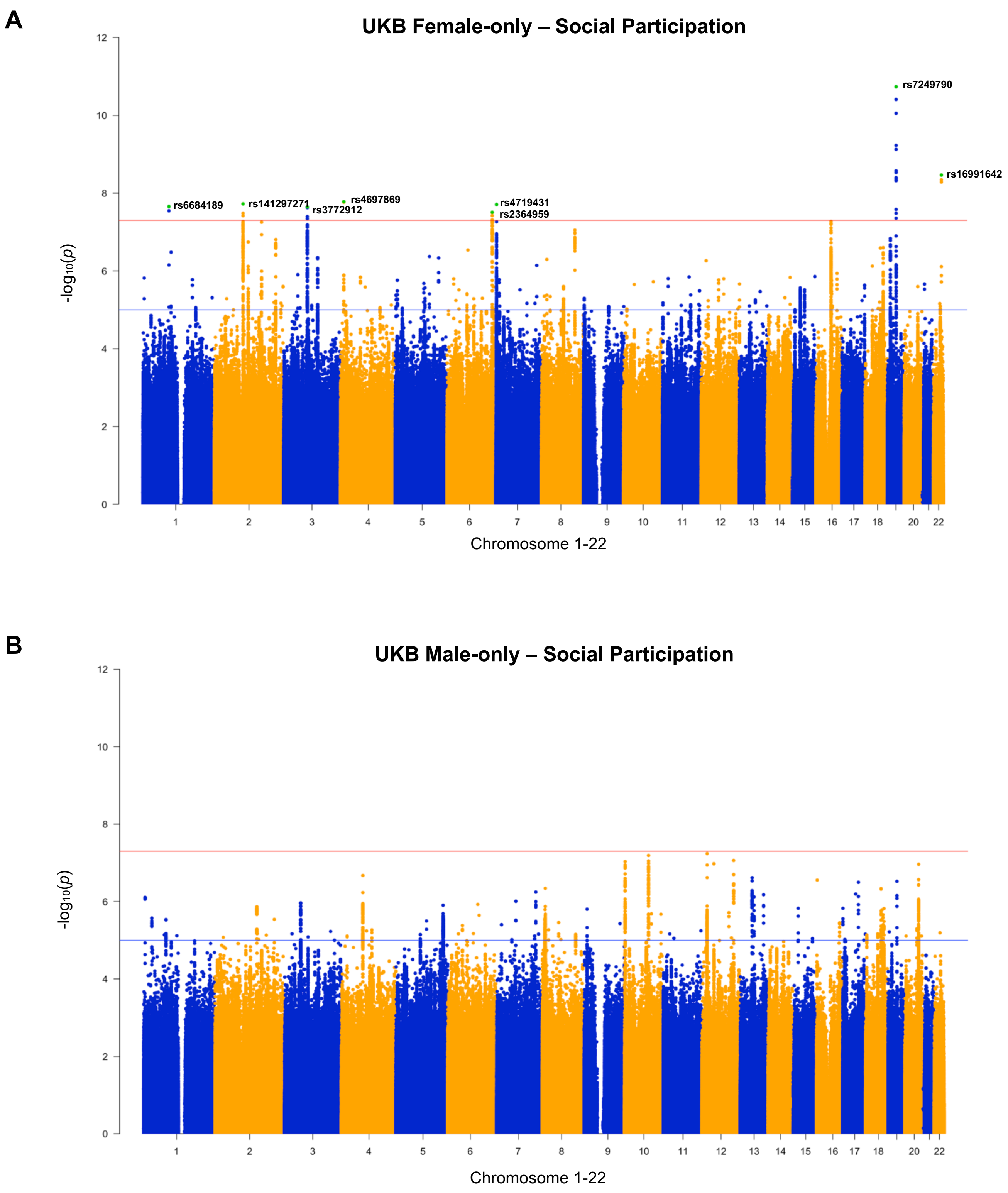


**Supplementary Figure 3 A** Manhattan plot of the observed -$\log_{10}$ *P*-values (*y*-axis) and the distribution of SNPs across chromosomes (*x*-axis) associated with the derived SP phenotype in the female-only cohort (*n*=218,757). The red line indicates the GWS threshold (*P*<5e-08). **B** Manhattan plot of the observed -$\log_{10}$ *P*-values for the SP phenotype in the male-only UK Biobank cohort (*n*=185,646). **Note**: Green denotes the lead variants identified.

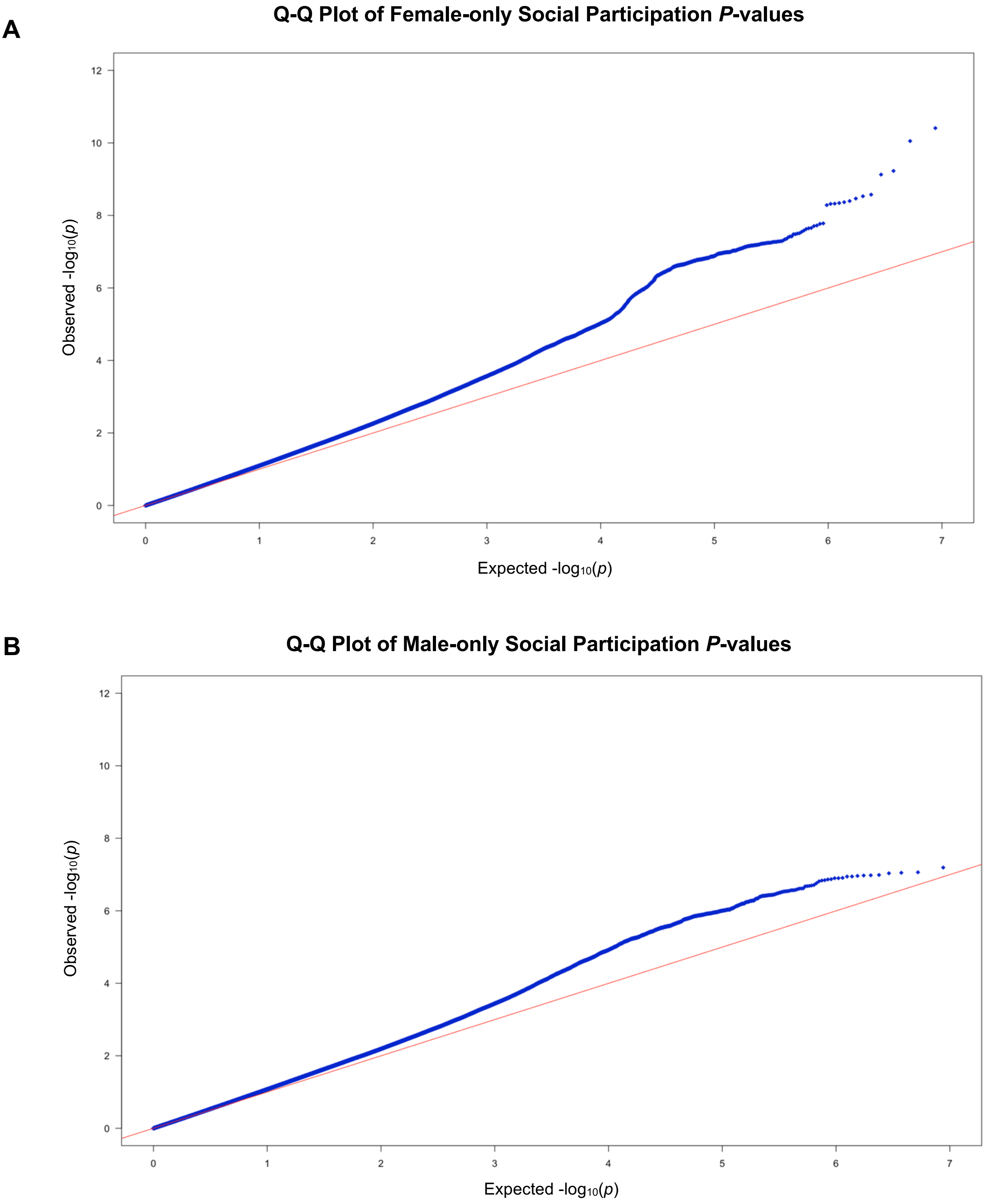


**Supplementary Figure 4** Q-Q plot of *P*-values for the SNP-based association analyses of the UKB SP phenotype in the **A** female-only sample and **B** male-only sample.

A

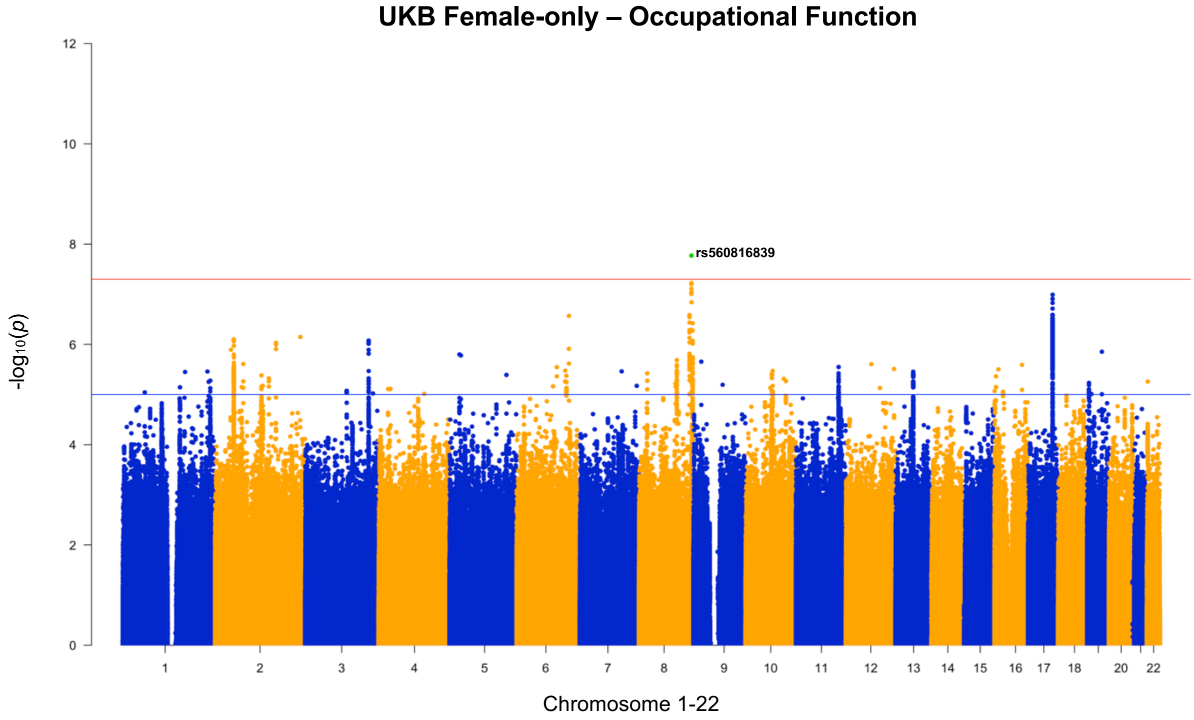


B

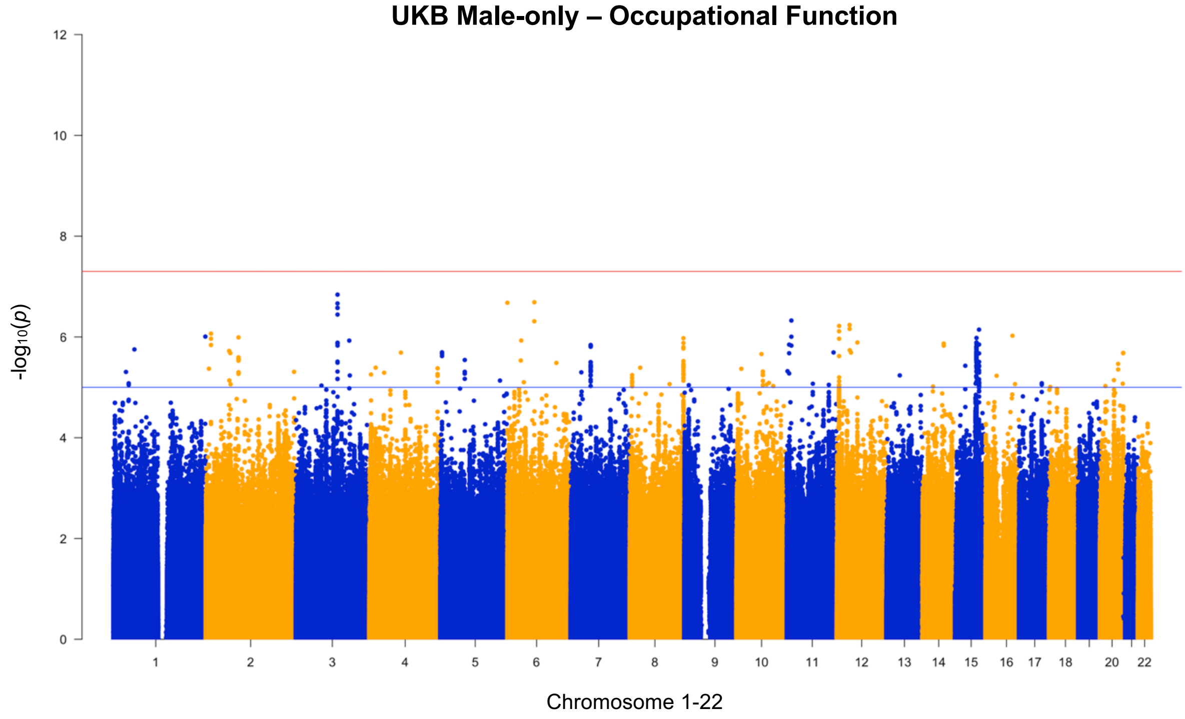


**Supplementary Figure 5 A** Manhattan plot of the observed -log$_{10}$ *P*-values (*y*-axis) and the distribution of SNPs across chromosomes (*x*-axis) associated with the derived OF phenotype in the female-only cohort (*n*=218,722). The red line indicates the GWS threshold (*P*<5e-08). **B** Manhattan plot of the observed -log$_{10}$ *P*-values for the OF phenotype in the male-only UK Biobank cohort (*n*=185,847). **Note**: Green denotes the lead variants identified.

**A**

**Q-Q Plot of Female-only Occupational Function *P*-values**

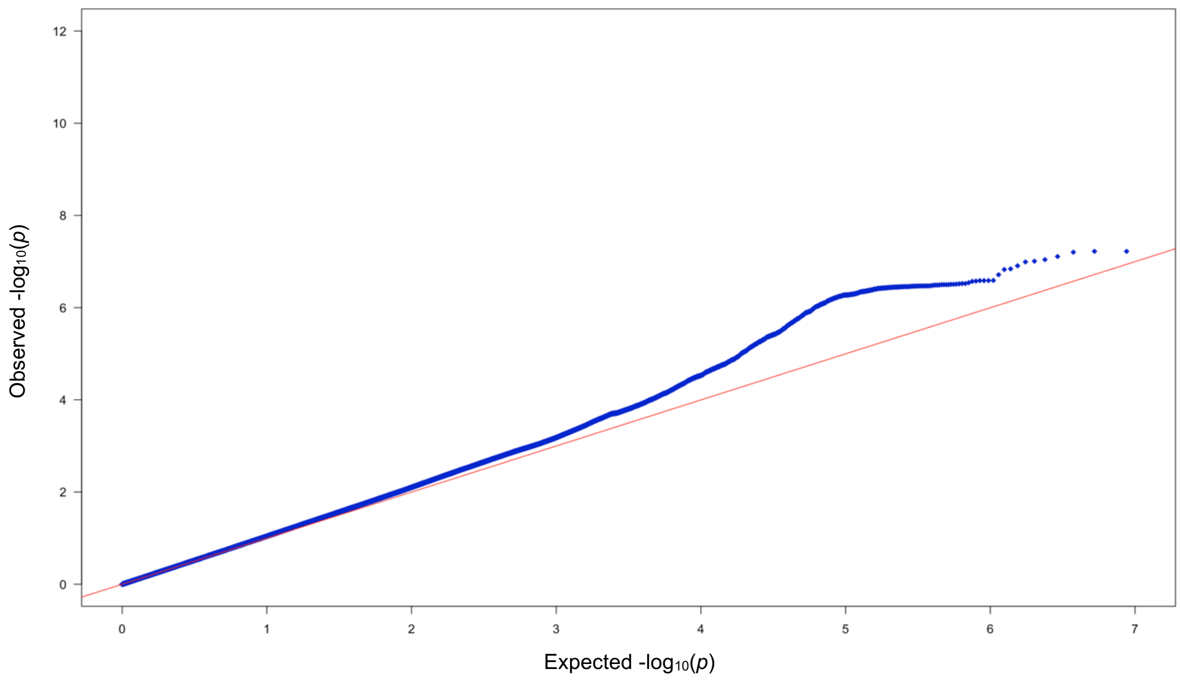


**B**

**Q-Q Plot of Male-only Occupational Function *P*-values**

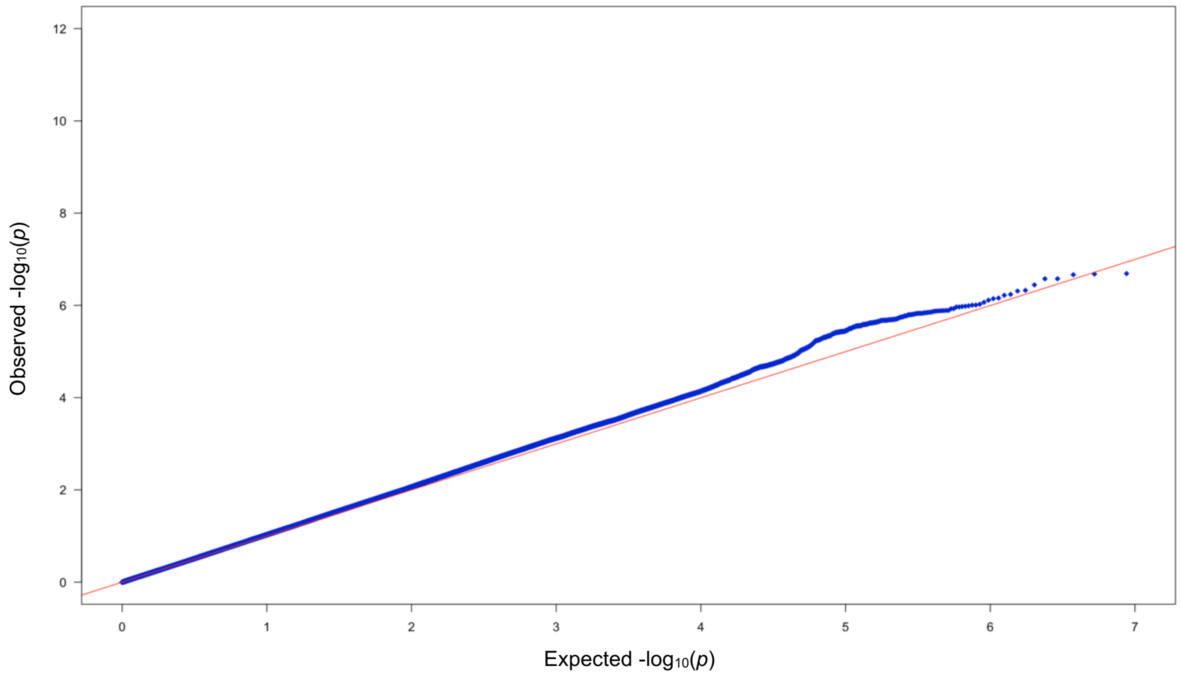


**Supplementary Figure 6** Q-Q plot of *P*-values for the SNP-based association analyses of the UKB OF phenotype in the **A** female-only sample and **B** male-only sample.

**A**

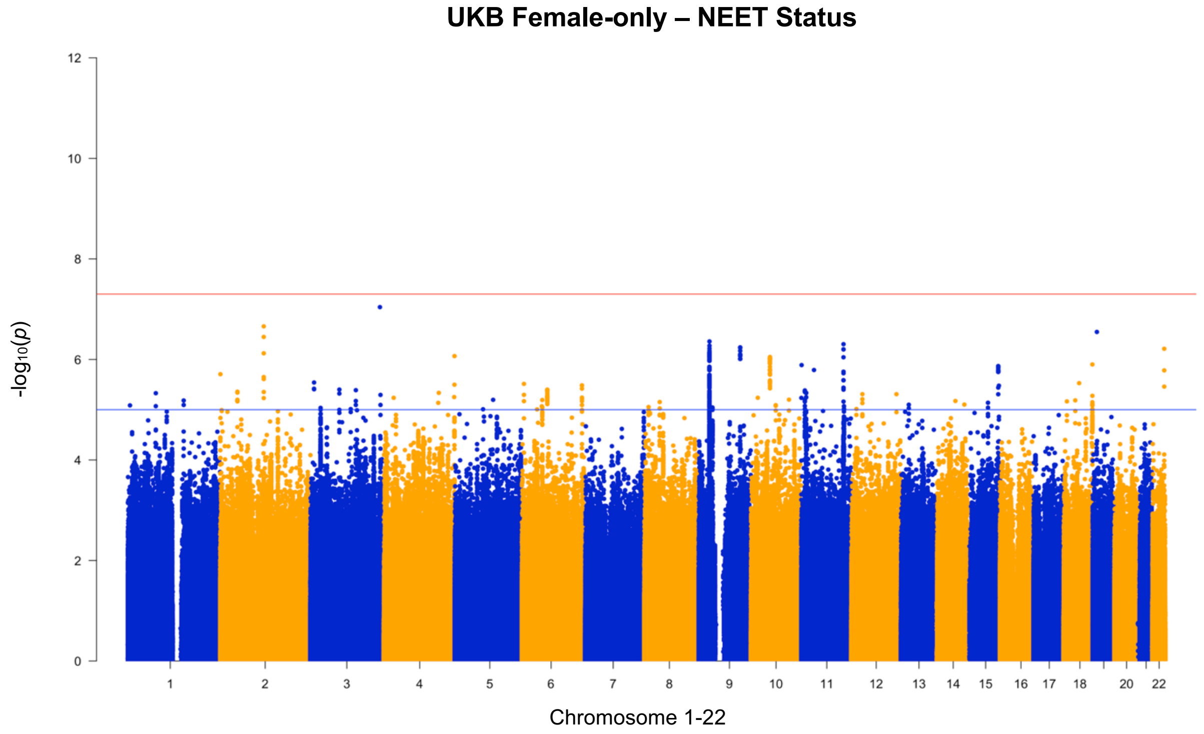


**B**

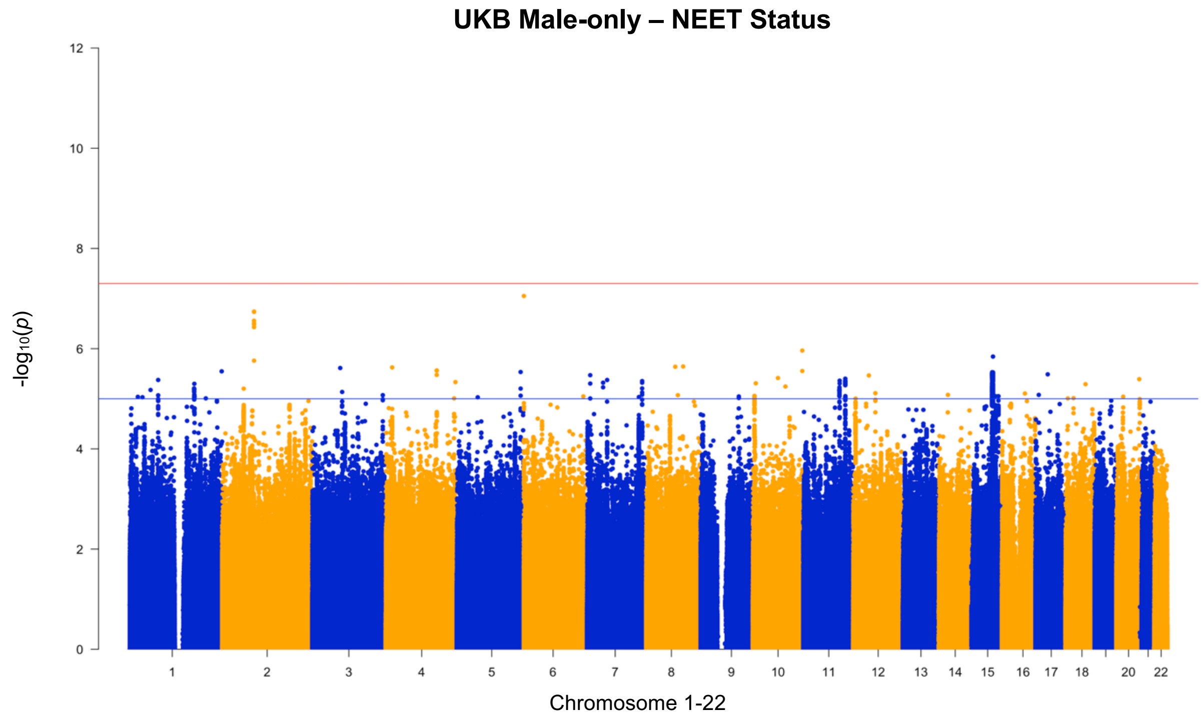


**Supplementary Figure 7 A** Manhattan plot of the observed -$\log_{10}$ *P*-values (*y*-axis) and the distribution of SNPs across chromosomes (*x*-axis) associated with the derived NEET status phenotype in the female-only cohort (*n*=218,686). The red line indicates the GWS threshold (*P*<5e-08). **B** Manhattan plot of the observed -$\log_{10}$ *P*-values for the NEET status phenotype in the male-only UK Biobank [illegible]

A

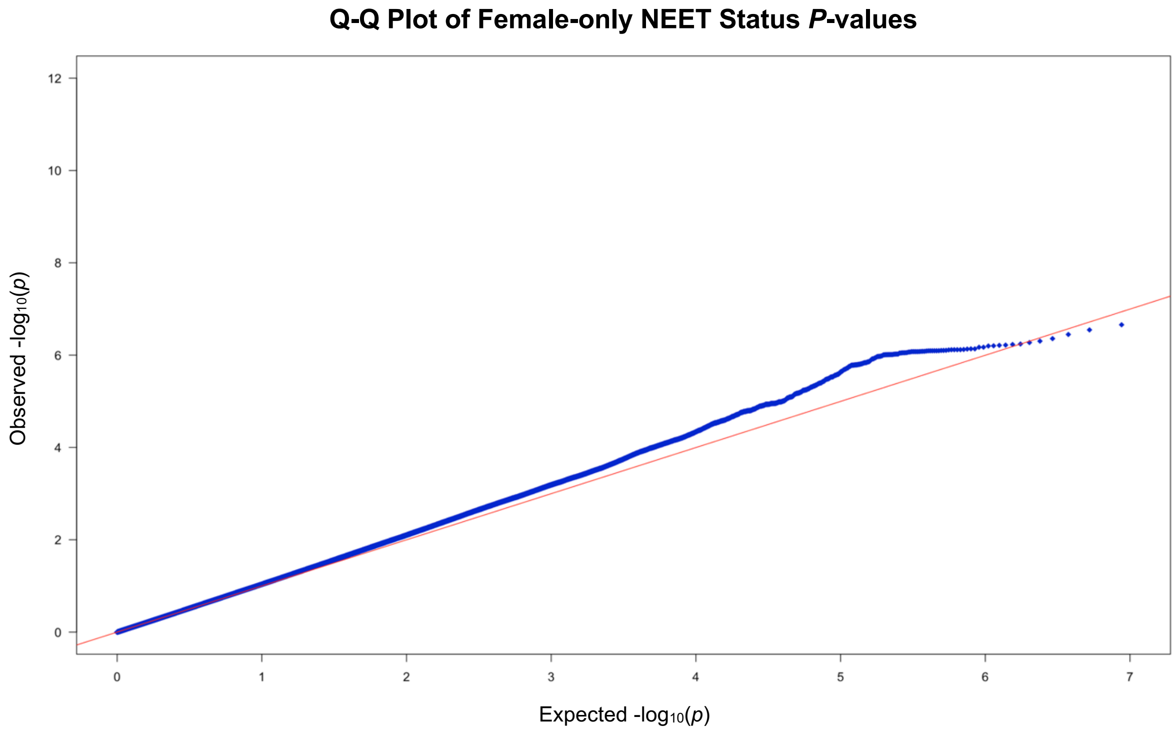


B

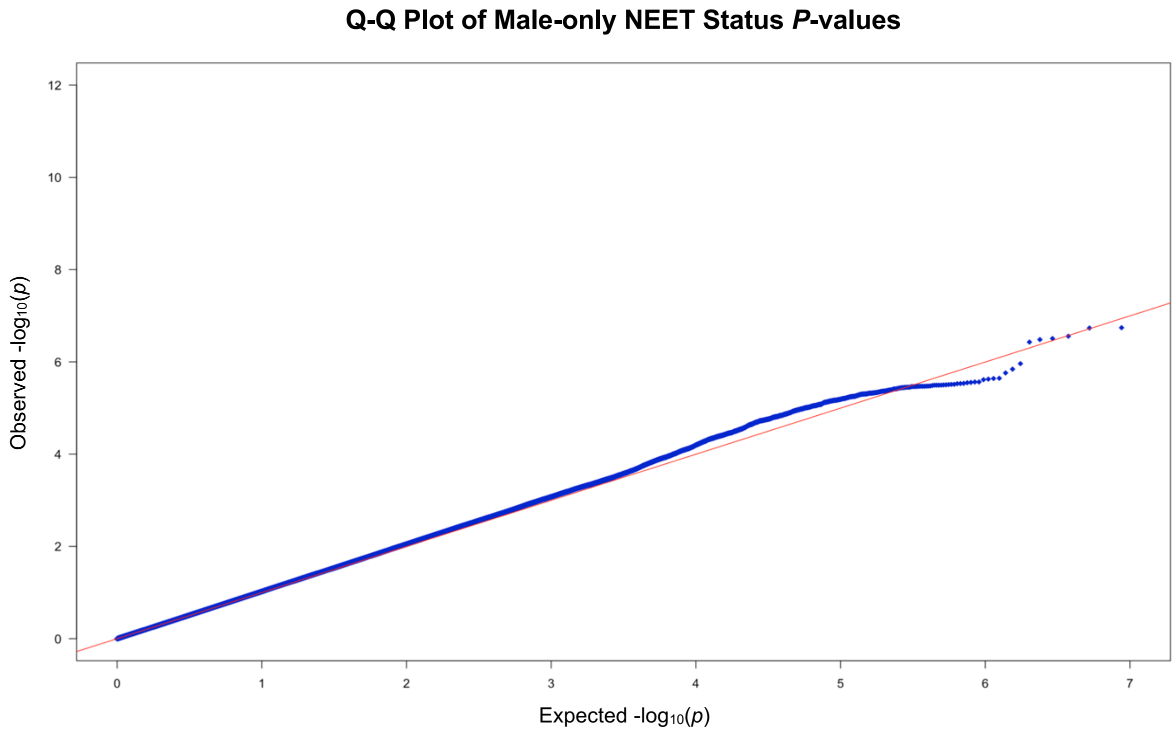


**Supplementary Figure 8** Q-Q plot of *P*-values for the SNP-based association analyses of the UKB NEET status phenotype in the **A** female-only sample and **B** male-only sample.

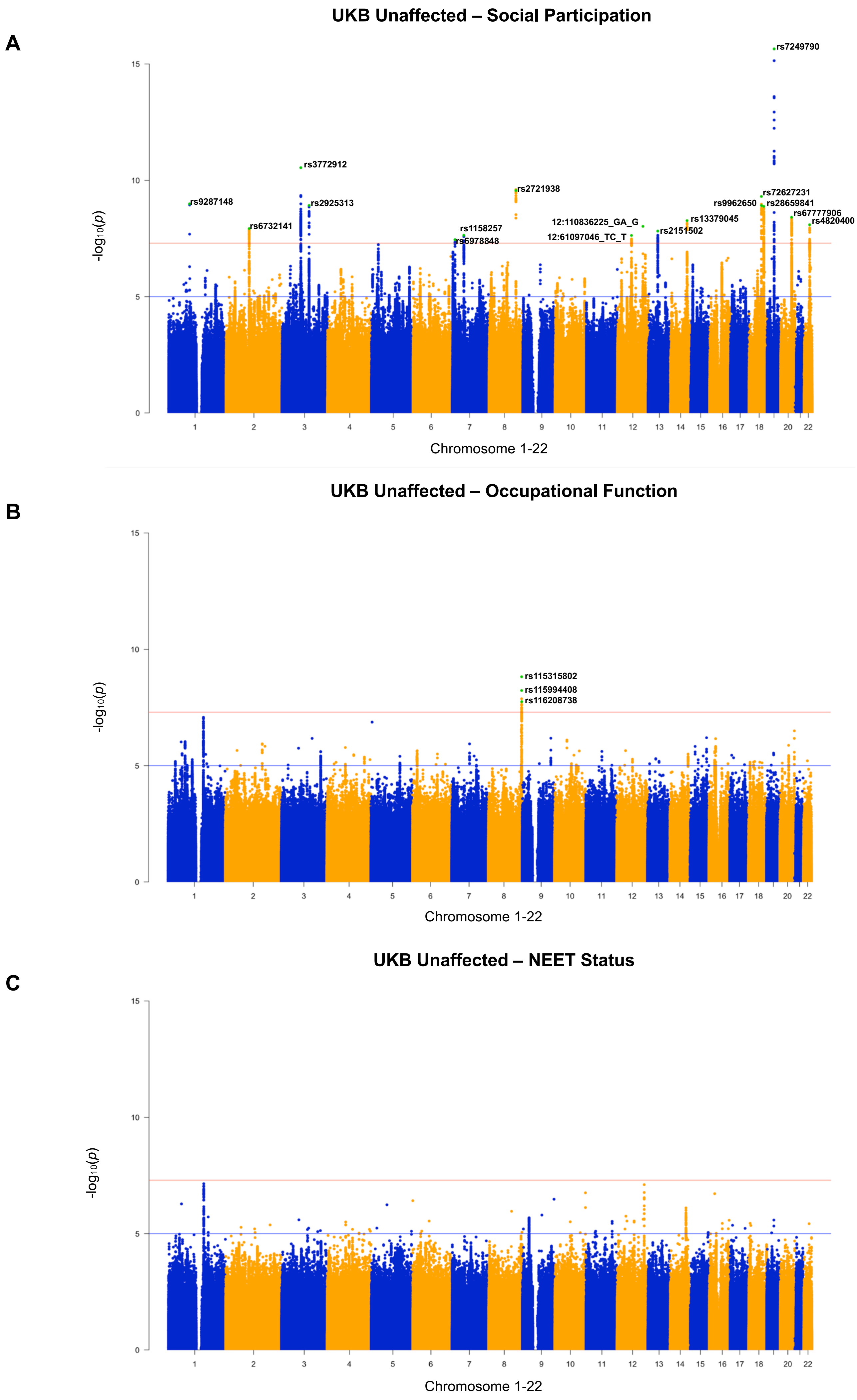
A
UKB Unaffected – Social Participation
rs7249790
rs3772912
rs2721938
rs9287148
rs2925313
rs6732141
rs1158257
rs6978848
12:110836225_GA_G
12:61097046_TC_T
rs2151502
rs13379045
rs9962650
rs72627231
rs28659841
rs67777906
rs4820400
-log10(p)
Chromosome 1-22
B
UKB Unaffected – Occupational Function
rs115315802
rs115994408
rs116208738
-log10(p)
Chromosome 1-22
C
UKB Unaffected – NEET Status
-log10(p)
Chromosome 1-22

**Supplementary Figure 9 A** Manhattan plot of the observed -$\log_{10}$ *P*-values (*y*-axis) and the distribution of SNPs across chromosomes (*x*-axis) associated with the derived SP variable in the UK Biobank unaffected cohort (*n* = 375,751). The red line indicates the GWS threshold (*P*<5e-08). **B** Manhattan plot of the observed -$\log_{10}$ *P*-values of the single nucleotide polymorphisms associated with the derived OF variable in the UK Biobank unaffected cohort (*n* = 375,856). **C** Manhattan plot of the observed -$\log_{10}$ *P*-values of the single nucleotide polymorphisms associated with the derived NEET status variable in the UK Biobank unaffected cohort (*n* = 375,869). **Note**: Green denotes the lead variants identified.

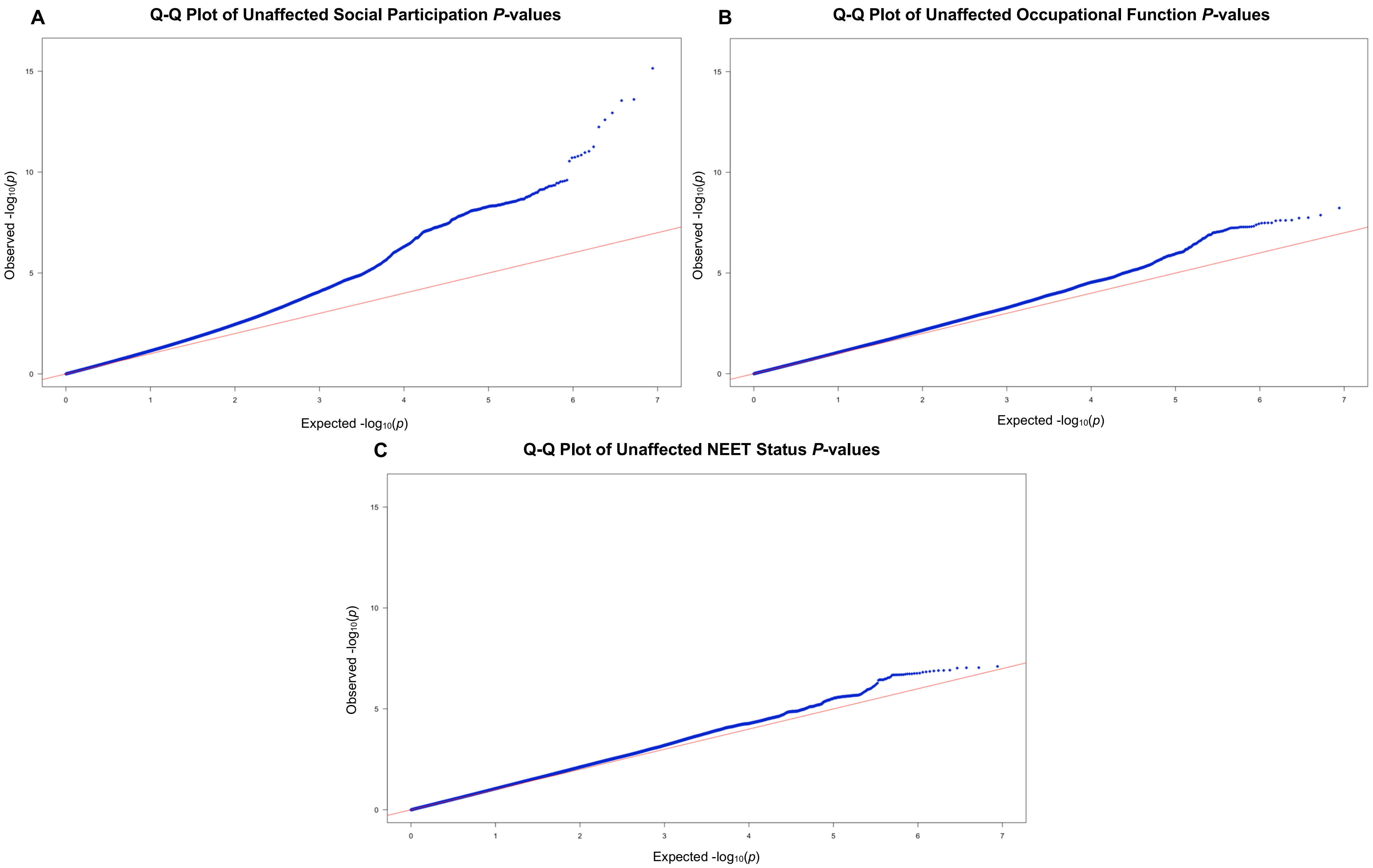


**Supplementary Figure 10** Q-Q plot of *P*-values for the SNP-based association analyses of the UKB phenotypes in the Unaffected Group: **A** SP, **B** OF, and **C** NEET status.

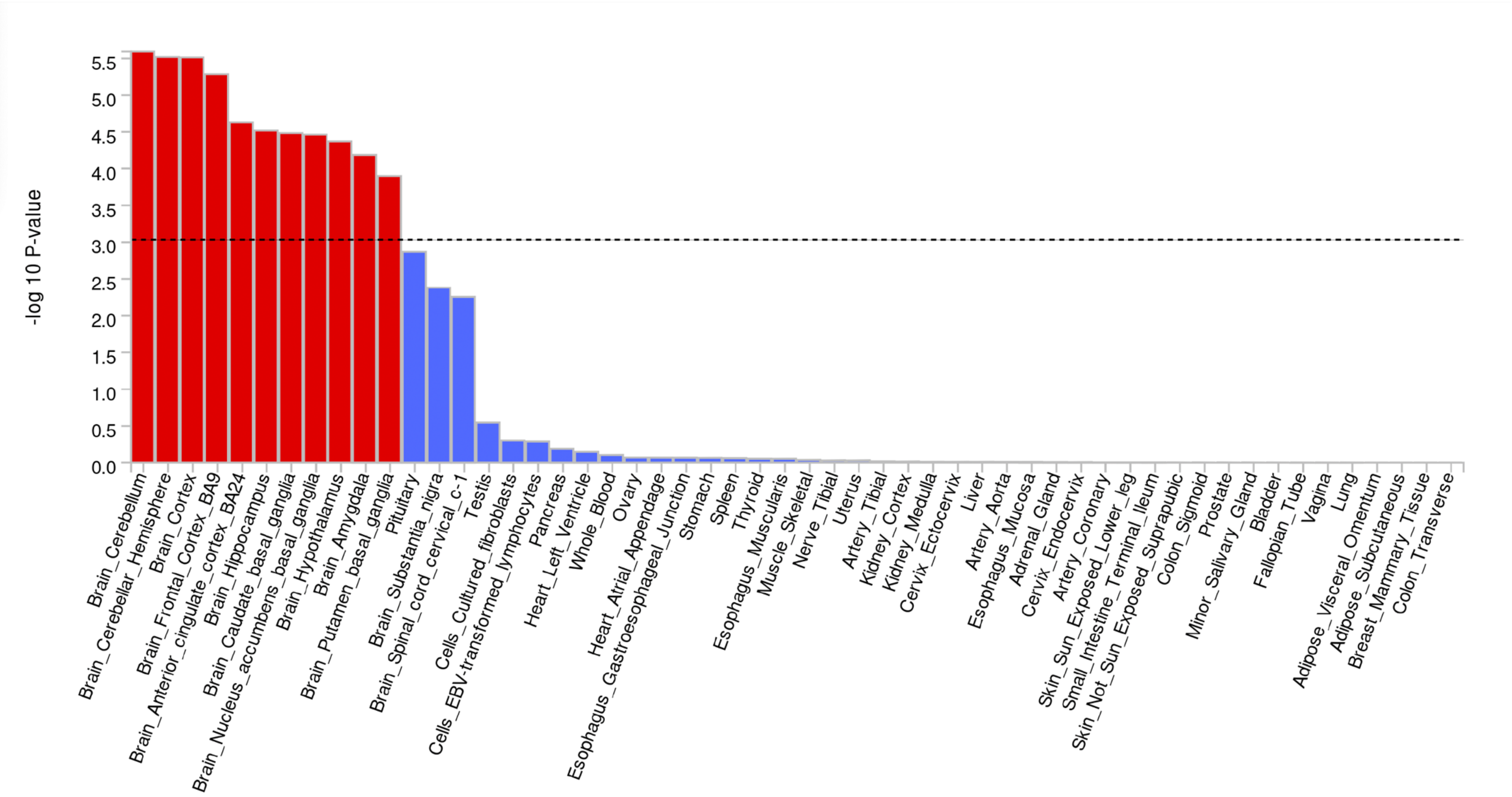


**Supplementary Figure 11** A bar plot depicting the various tissue types within which the SP-related genes were most significantly enriched. The x-axis shows the names of the analysed tissues, and the y-axis shows the $\log_{10}$ *P*-values associated with each tissue type. Red bars indicate a significant enrichment.

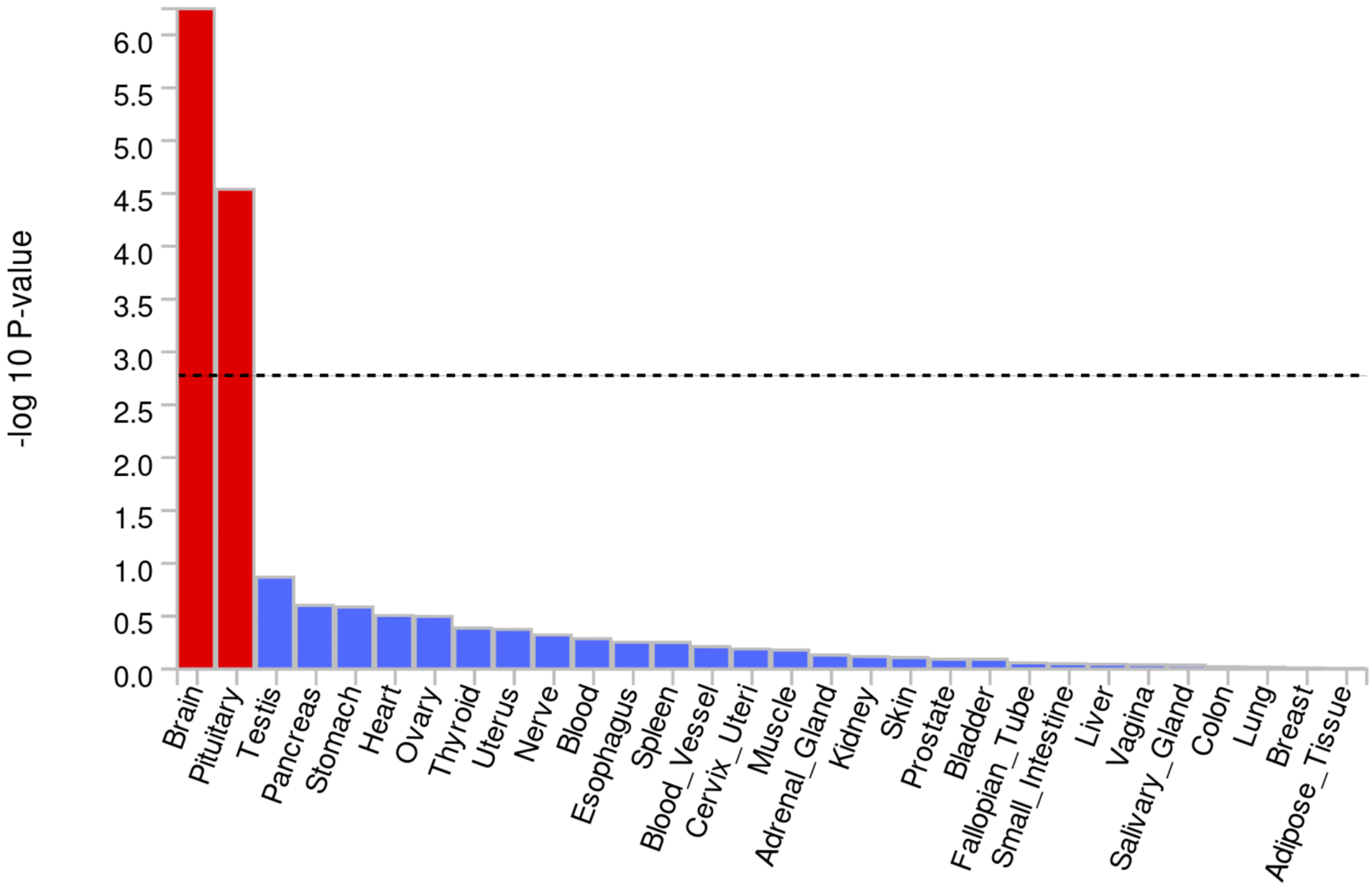


**Supplementary Figure 12** A bar plot depicting the general tissue types within which the SP-related genes were most significantly enriched. The x-axis shows the names of the analysed tissues, and the y-axis shows the $\log_{10}$ *P*-values associated with each tissue type. Red bars indicate a significant enrichment.

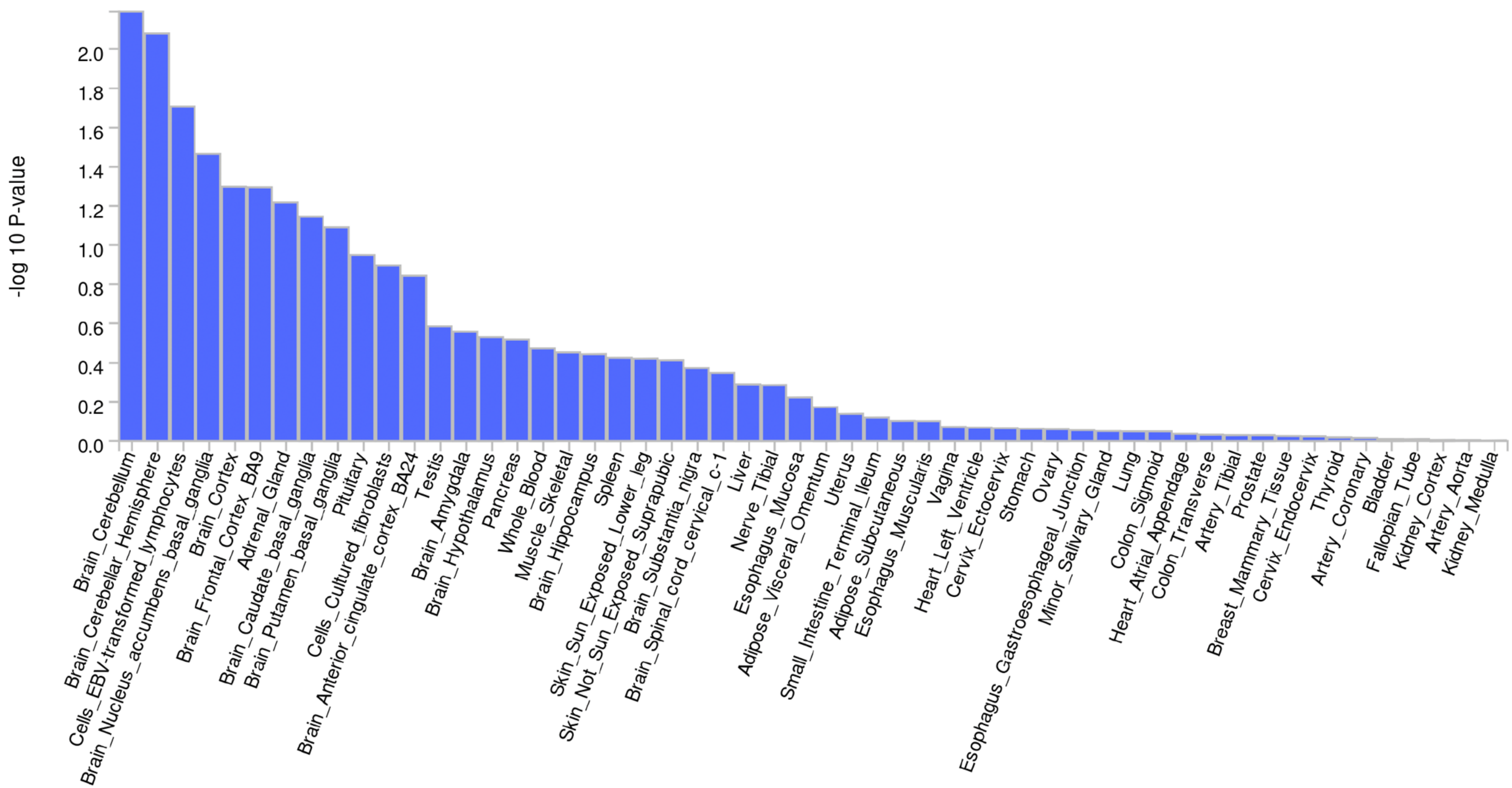


**Supplementary Figure 13** A bar plot depicting the various tissue types within which the OF-related genes were most significantly enriched. The x-axis shows the names of the analysed tissues, and the y-axis shows the $\log_{10}$ *P*-values associated with each tissue type. The absence of red bars indicates that there was no significant enrichment within a specific tissue type.

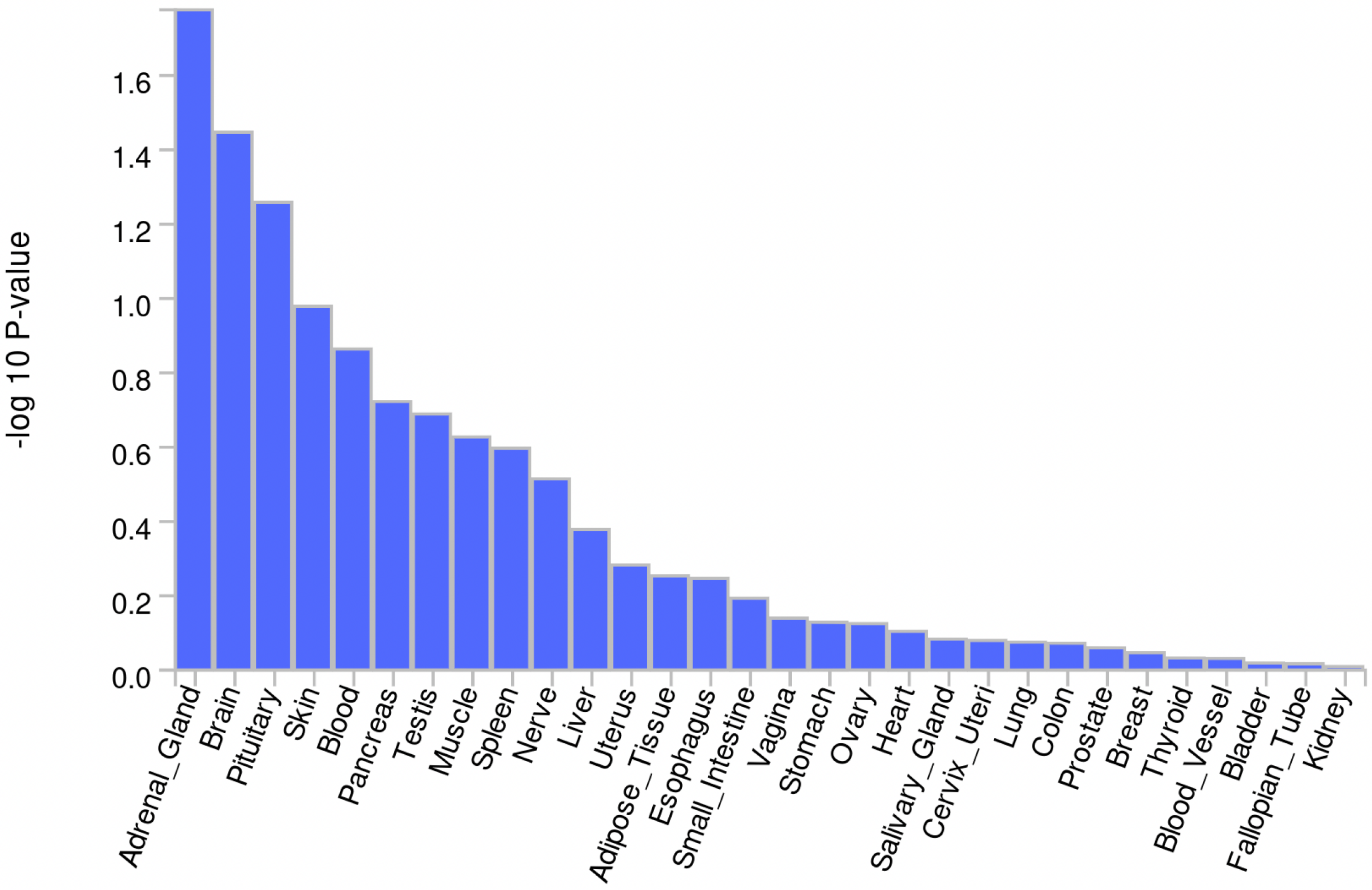


**Supplementary Figure 14** A bar plot depicting the general tissue types within which the OF-related genes were most significantly enriched. The x-axis shows the names of the analysed tissues, and the y-axis shows the $\log_{10}$ *P*-values associated with each tissue type. The absence of red bars indicates that there was no significant enrichment within a specific tissue type.

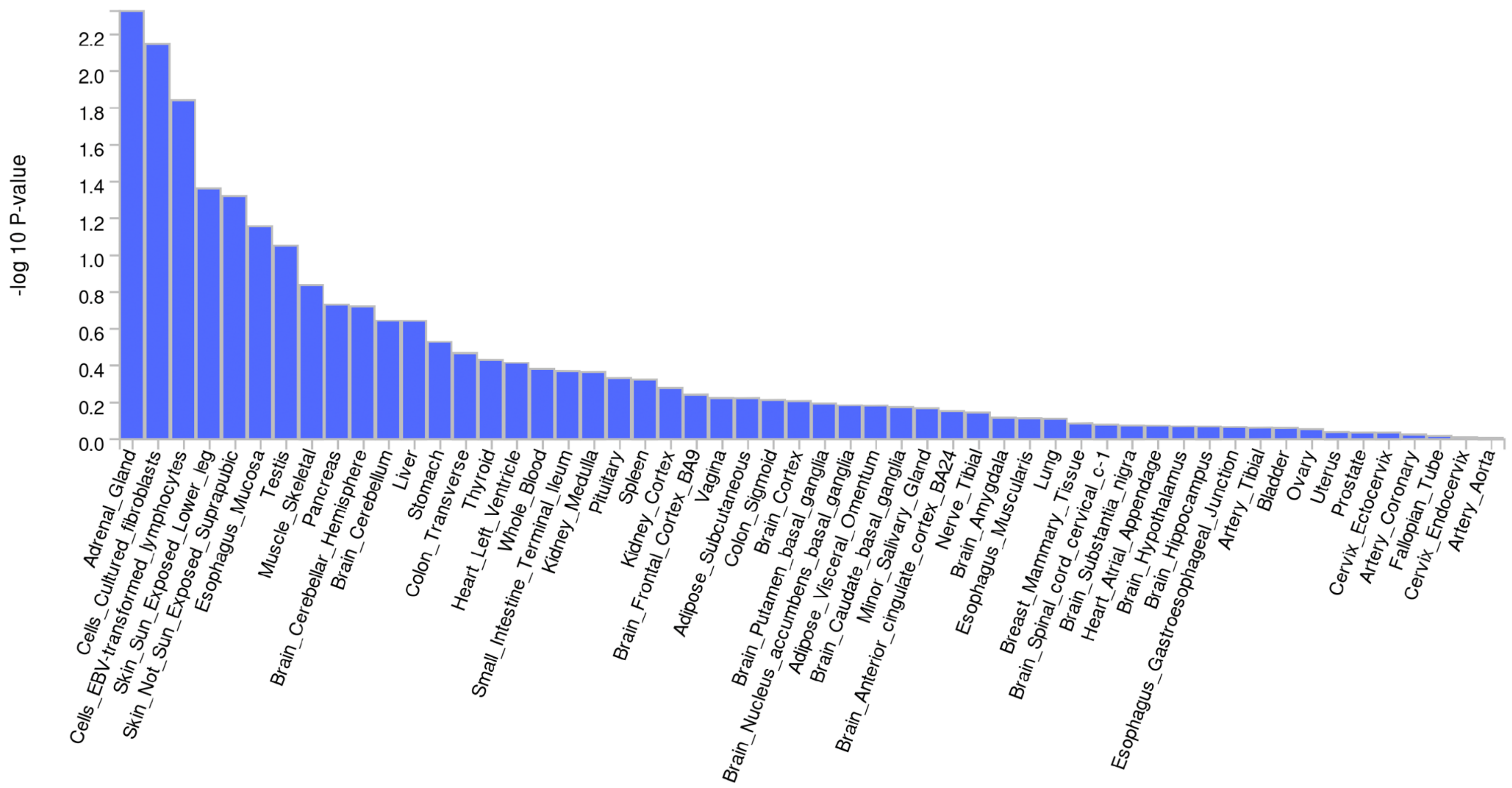


**Supplementary Figure 15** A bar plot depicting the various tissue types within which the NEET status-related genes were most significantly enriched. The x-axis shows the names of the analysed tissues, and the y-axis shows the $\log_{10}$ *P*-values associated with each tissue type. The absence of red bars indicates that there was no significant enrichment within a specific tissue type.

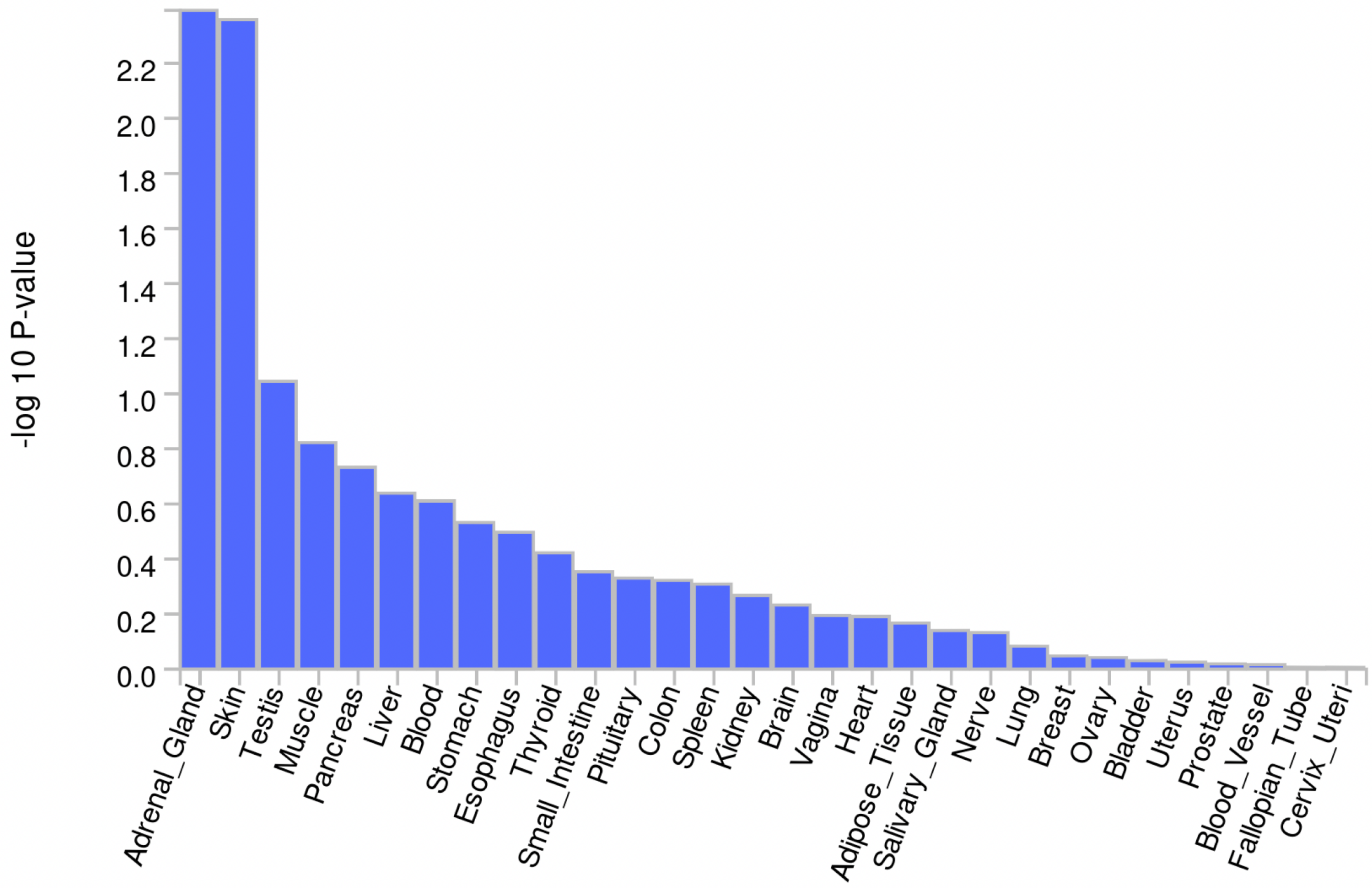


**Supplementary Figure 16** A bar plot depicting the general tissue types within which the NEET status-related genes were most significantly enriched. The x-axis shows the names of the analysed tissues, and the y-axis shows the $\log_{10}$ *P*-values associated with each tissue type. The absence of red bars indicates that there was no significant enrichment within a specific tissue type.

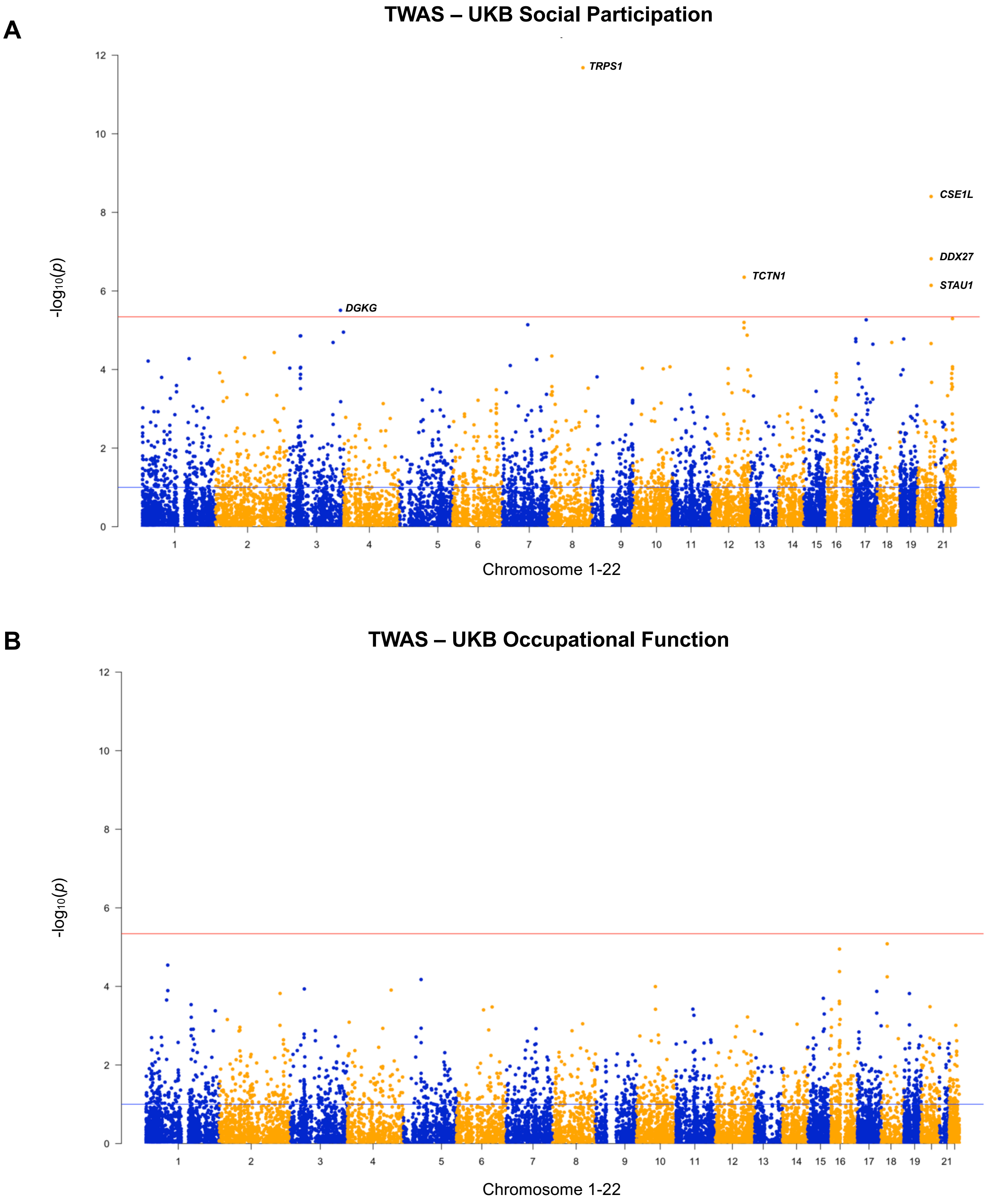


**Supplementary Figure 17 A** Manhattan plot of the observed -log$_{10}$ *P*-values (*y*-axis) and the distribution of genes (*N*=11,934 genes) across chromosomes 1-22 (*x*-axis) associated with the derived SP phenotype. The red line indicates the GWS threshold (*P*<4.19e-06); **B** Manhattan plot of the observed -log$_{10}$ *P*-values (*y*-axis) and the distribution of genes (*N*=11,934 genes) across chromosomes 1-22 (*x*-axis) associated with the derived OF phenotype.
